# A “Tail” of Two Cities: Fatality-based Modeling of COVID-19 Evolution in New York City and Cook County, IL

**DOI:** 10.1101/2020.08.10.20170506

**Authors:** Joshua A. Frieman

## Abstract

I describe *SIR* modeling of the COVID-19 pandemic in two U.S. urban environments, New York City (NYC) and Cook County, IL, from onset through mid-June, 2020. Since testing was not widespread early in the pandemic in the U.S., I rely on public fatality data to estimate model parameters and use case data only as a lower bound. Fits to the first 20 days of data determine a degenerate combination of the basic reproduction number, *R*_0_, and the mean time to removal from the infectious population, *γ*^−1^, with *γ*(*R*_0_ − 1)= 0.25(0.21) inverse days for NYC (Cook County). Equivalently, the initial doubling time was *t_d_* = 2.8(3.4) days for NYC (Cook). The early fatality data suggest that both locations had infections in early February. I model the mitigation measures implemented in mid-March in both locations (distancing, quarantine, isolation, etc) via a time-dependent reproduction number *R_t_* that declines monotonically from *R*_0_ to a smaller asymptotic value, *R*_0_(1 − *X*), with a parameterized functional form. The timing (mid-March) and duration (several days) of the transitions in *R_t_* appear well determined by the data. With flat priors on model parameters and the lower bound from reported cases, the NYC fatality data imply 95.45% credible intervals of *R*_0_ = 2.6 − 2.9, social contact reduction *X* = 69 − 76% and infection fatality rate *f* = 1 − 1.5%, with 19 − 27% of the population asymptotically infected. The case data relative to daily deaths suggest that the reported case rate as a fraction of true case rate grew monotonically, reaching a plateau around April 20 for both NYC and Cook County; the models also suggest that the late-time NYC reported case rate was comparable to the true rate, while for Cook County it remained an underestimate. For Cook County, the fatality evolution was qualitatively different from NYC: after mitigation measures were implemented, daily fatality counts reached a plateau for about a month before tailing off. This is consistent with an *SIR* model that exhibits “critical slowing-down”, in which *R_t_* plateaus at a value just above unity. For Cook County, the 95.45% credible intervals for the model parameters are not constrained by the case data and are much broader, *R*_0_ = 1.4 − 4.7, *X* = 26 − 54%, and *f* = 0.1 − 0.6% with 15 − 88% of the population asymptotically infected. Despite the apparently lower efficacy of its social contact reduction measures, Cook County has had significantly fewer fatalities per population than NYC, *D*_∞_ /*N* = 100 vs. 270 per 100,000. In the model, this is attributed to the lower inferred IFR for Cook; an external prior pointing to similar values of the IFR for the two locations would instead chalk up the difference in *D*/*N* to differences in the relative growth rate of the disease. I derive a model-dependent threshold, 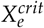, for ‘safe’ re-opening, that is, for easing of contact reduction that would not trigger a second wave; for NYC, the models predict that increasing social contact by more than 20% from post-mitigation levels will lead to renewed spread, while for Cook County the threshold value is very uncertain, given the parameter degeneracies. The timing of 2nd-wave growth will depend on the amplitude of contact increase relative to 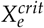 and on the asymptotic growth rate, and the impact in terms of fatalities will depend on the parameter *f*.

## 1 Introduction

Epidemiological models of infectious diseases can be useful in inferring characteristics such as disease transmissibility within a given population and for modeling disease progression, dependencies, and the impact of mitigation measures. Typically, models are fit retrospectively to historical disease data (e.g., Ebola [1] and previous coronavirus outbreaks). At early stages of the COVID-19 pandemic, public data sets were used to infer some characteristics of the disease in different regions (e.g., [2-3]), but forecasting its future progression is a fraught exercise at best: the data are incomplete and biased due to incomplete and non-random sampling, the models are overly simplistic in their dynamics and assumptions, and public health measures are constantly changing and highly heterogeneous. Nevertheless, models can be used to make ‘what-if’ projections that describe an envelope of outcomes given current uncertainties and show the impact of taking or not taking timely mitigation measures [4]. In addition, exploring simple dynamical models can determine if few-parameter models have enough complexity to reasonably describe the data and, if so, to draw conclusions about model parameters describing the pandemic.

In this note, I present a comparative study of COVID-19 evolution in 2 U.S. urban environments, New York City and Chicago and environs, with the goal of exploring their similarities and differences and seeing what qualitative conclusions might be drawn at this stage, using the simplest version of the Susceptible-Infected-Recovered (*SIR*) epidemiological model. As of this writing (mid-July), both locations have experienced hundreds of thousands of COVID-19 infections and thousands of deaths, but the ‘first wave’ of the pandemic has largely run its course, with daily fatalities in both locations now in the single digits. The time is ripe to retrospectively study this phase and see what lessons might be drawn for the next. To determine model parameters, I primarily use public fatality data, since the case data are subject to much larger selection effects; however, I do further constrain parameters by employing case data as a lower bound on true cases.

## 2 Susceptible-Infected-Recovered (SIR) Model

The *SIR* model is a simplified version of a more general infectious disease transmission model studied by Kermack and McKendrick (and predecessors) in the late 1920’s and early 1930’s [5]. Similar to the Lotka-Volterra model for the predator-prey problem introduced a few years earlier, it relies on a system of coupled, first-order differential equations to describe the interactions of and transformations between subpopulations. In the form used here, the *SIR* model describes only mean community transmission in a population once infection is present; it does not account for diffusion within a community nor spatial spread between communities in different regions. It can be thought of as a kind of mean-field approximation to more detailed granular dynamics [6].

The SIR model assumes a “closed”, spatially homogeneous, interacting population or community of *N* individuals, comprising *S* susceptible, *I* infected/infectious, and *R* recovered/removed individuals, so that

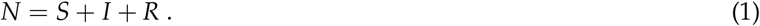

More complex versions of the model include additional subpopulations (e.g., exposed, latent, symptomatic, hospitalized, etc), with associated additional parameters. A susceptible (non-infected) individual interacting with other individuals has an average daily probability of getting infected of *p_i_* = *βI*/*N*, where *I*/*N* is the fraction of the population currently infected, and *β* is a characteristic parameter with units of inverse time that encodes the transmissability of the disease and the typical number of daily encounters per susceptible individual. Due to these transmissive encounters, the susceptible population, *S*, declines according to

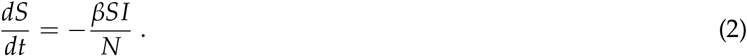

Defining *γ*^−1^ as the mean time in days for an infected individual to be removed from the infectious population, either due to recovery, hospitalization, isolation/quarantine, or death, the evolution of the infected population, *I*, is given by

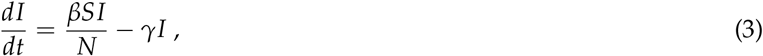

where the first term on the RHS of Eqn. (3) follows from Eqn. (2). By conservation of individuals, the rate of change of the number of removed/recovered individuals, *R*, must satisfy

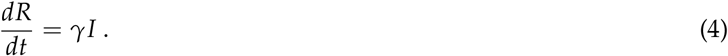

Note that this version of the model refers only to those infected, whether or not they display illness symptoms, and recovered/removed just means no longer contagious and interacting in the population, again independent of illness symptoms. It is also assumed here that recovery confers immunity on a timescale longer than any of those considered here.

The Infection Fatality Rate, *f*, is taken to be the ratio of the asymptotic, cumulative number of those who die from the disease, *D*(*t* = ∞), to the number of individuals ever infected, *f* = *D*(*t* = ∞)/*R*(*t* = ∞), where “asymptotic” implies post-pandemic, that is, *I*(*t* = ∞)= 0. In the modeling that follows, I further assume that *f* is a fixed (time-independent) parameter for a given community. For finite times, since there is a time delay between infection and fatality, I allow for a time-translation, viz.,

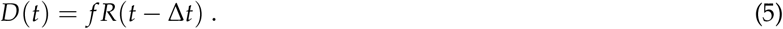

Since there is clear evidence that the subpopulation of those who have died from COVID-19 is not a random subset of the infected population and that improvements in hospital care over time have improved disease outcomes, the assumption that *f* remains time-independent within a community should be viewed with skepticism. For the analysis here, I am making the weaker (but still questionable) assumption that the timescale for *f* to vary has, so far, been long compared to the 3.5-month period under study.

The *SIR* model is thus specified by the 3 dynamical equations (2), (3), and (4), and the constraint (1), so it is a dynamical system with 2 state variables. Solutions are determined by the two dimensionful parameters *β* and *γ*, the dimensionless parameter *f* (which only enters when comparing to fatality data), and a specified initial condition at time *t*_0_, which is conventionally taken as the initial number of cases, *I*(*t*_0_) ≡ *I*_0_, with, from Eqn. (1), *S*(*t*_0_)= *N* − *I*_0_, assuming *R*(*t*_0_)= 0 at sufficiently early time. The transmission rate, *β*, is expected to depend on community characteristics (population density, mobility, modes of transportation, mitigation measures in place, etc,). By contrast, the mean removal time, *γ*^−1^, is determined to a degree by the human body’s response to the disease, so it might be expected to be more of an “intrinsic” property that would vary less from community to community, although it is certainly affected by variations in age, health, and customs between communities, as well as by public awareness, the availability of testing, etc.

### 2.1 Scaled Version of the Model

The dynamics can be made more transparent by defining scaled variables for the fractions of susceptible, infected, and removed, *s* = *S*/*N*, *i* = *I*/*N*, *r* = *R*/*N*, a dimensionless time variable, *τ* = *γt*, and the dimensionless *basic reproduction number*, *R*_0_ = *β*/*γ*, in terms of which the model equations become

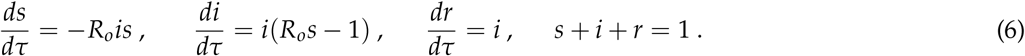

The scaled dynamics are determined completely by *R*_0_ and by the initial condition, i.e., by the value of *i*_0_ = *I*_0_/*N*. The infection will initially grow if *R*_0_ > 1. Since *D* = *f Nr*, for a given model (*r*) and fatality data set (*D*), the parameters *f* and *N* are perfectly degenerate.

Early on, when only a tiny fraction of the population is infected, *s* ≃ 1 − *I*_0_ ≃ 1, the growth is quasi-exponential,

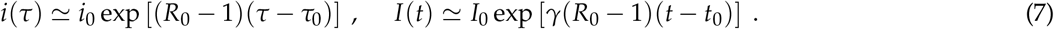

In this limit, the dimensionless doubling time of infections in the population is *τ_d_* = ln 2/(*R*_0_ − 1)= 0.7/(*R*_0_ − 1) and therefore *t_d_* = 0.7/*γ*(*R*_0_ − 1) days. As the infection spreads and *s* starts to decline, the infection growth rate *di*/*dτ* slows, and the number of infected reaches a maximum (*di*/*dτ* = 0) when *s* = 1/*R*_0_ and thereafter declines. Alternatively, the number of infected will decline when *R*_0_ < 1/*s*. The daily death rate, *dD*/*dt*, peaks a time Δ*t* later. The scaled daily rate of new cases, *dc*/*dτ* = *R*_0_*is*, peaks when *d*(*is*)/*dτ* = 0; using Eqn. (6), this occurs when *s* − *i* = 1/*R*_0_. Since *s* is monotonically falling, the rate of new cases peaks earlier than the peak in number of infected. An obvious point is that the asymptotic future value of *s* must be less than 1/*R*_0_, so the asymptotic future value of *r* satsifies *r*_∞_ > 1 − 1/*R*_0_, assuming that the disease does not remain endemic (*i* → 0 asymptotically): a substantial fraction of the population will get infected unless *R*_0_ is very close to 1. For example, for *R*_0_ = 3, about *r*_∞_ = 94% of the population will get infected or else 2/3 of the population would need to be vaccinated to stop the spread. All of these statements assume that no mitigation measures are taken, so that *R*_0_ is constant in time. Since *R*_0_ is typically of order unity, we expect *a priori* that the dynamics will yield quantities of order unity as well; however, the initial condition *i*_0_ ≪ 1 provides a very small dimensionless parameter that can lead to large values for the characteristic dynamical time *τ* ≫ 1.

It’s useful to relate the *SIR* model parameters to characteristic quantities related to transmission of the infection. The probability per day that an infected individual transmits infection to a susceptible individual is *p_t_* = *βS*/*N* = *R*_0_*γs* ≃ *R*_0_*γ* at the beginning of the pandemic. Assuming the probability of infecting another individual is a Poisson process with rate *λ* = *pt*, then early in the pandemic the mean number of individuals that an infected individual infects before removal from the population is *λ*/*γ* ≃ *R*_0_, since 1/*γ* is the mean time to removal. This is often taken as an operational definition of *R*_0_. For an individual infected at *t* = 0, if *t_a_* is the time until they first infect another individual and *t_s_* is the time interval between when they infect their first and second individuals, then for a Poisson process *t_a_* and *t_s_* both follow an exponential distribution, *p*(*t_i_*)= *λ* exp(−*λt_i_*), with mean 〈*t_i_*〉 = 1/*λ* and variance *σ*^2^(*t_i_*)= 1/*λ*^2^. Thus, the *serial time*, the mean time interval between an infected person’s first and second transmissions to other individuals, is 〈*t_s_*〉 = 1/*R*_0_*γs*. The infection will spread rapidly if the serial time is less than the removal time, 〈*t_s_*〉 < 1/*γ*, that is, if *R*_0_*s* > 1, in agreement with Eqn. (6).

### 2.2 SIR Model with Reduction of Social Contact

A variety of physical distancing and isolation measures were taken at the local, state, and national levels starting around mid-March that suppressed the growth of COVID-19 cases and thus of fatalities. In the epidemiology literature, such mitigation measures are often modeled by assuming that the basic reproduction number *R*_0_ in Eqn. (6) is replaced by a function *R_t_* that decays in time to a smaller final value [1]. This introduces 3 additional model parameters: the characteristic time when measures are taken, *t_m_*; the percentage reduction *X* in social contact, i.e., in *R_t_*, from well before to well after *t_m_*; and the characteristic time *α*^−1^ that it takes for the measures to go into effect, that is, the duration of the transition in *R_t_* from *R*_0_ to its final value of *R*_∞_ = *R*_0_(1 − *X*).

I have tried both exponential and hyperbolic tangent functions of time to model *R_t_*,

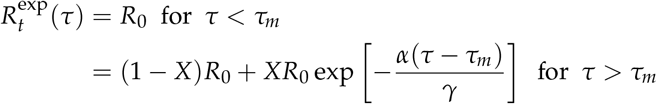

or

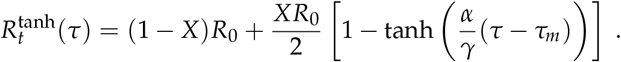

Note that for the tanh model, *t_m_* = *τ_m_*/*γ* is not the date of onset of mitigation measures but rather the midpoint of the transition in *R_t_* from *R*_0_ to *R*_∞_. I present results below for the tanh model, but the parameter inferences do not seem strongly dependent on which functional form is assumed. An alternative approach to adopting a parameterized functional form for *R_t_* would be to infer the model functions *s*, *i* and *r* empirically from the fatality data, up to the unknown model parameters *f* and *γ*; this is discussed in Appendix B. For the rest of the paper, I adopt and test the functional form of Eqn. (8) since unlike the non-parametric model it allows one to make parameter inferences and forecasts.

## 3 Estimating *R*_0_ from early Fatality Data

Due to the reported lack of deployment of test kits in the U.S. early in the pandemic, the raw public data on COVID-19 confirmed cases vs. time underestimated the true number of infections by a large, uncertain, and *time-dependent* factor. In many parts of the country, those with COVID-19-like symptoms who did not require hospitalization were not routinely tested, especially early on. Some early estimates were that only 10-20% of those infected showed severe symptoms that might prompt hospitalization, suggesting that the number of early infections may have been underestimated by a factor of 5-10. Time-dependence of this sampling incompleteness will introduce a spurious time-dependence in the confirmed-case rate and thus bias estimates of *R*_0_. In principle, this bias can be corrected using information on the (time-dependent) number of tests given per day, but given the very low sampling rate early on and the fact that testing for the most part has not been done on random subsamples of the population, the uncertainty in the bias correction factor could be significant.

For these reasons, I do not use data on the number of confirmed cases vs. time for estimating the SIR model parameters. Instead, I rely upon public fatality data, using reported cases only as a lower bound on the number of true cases. Recent studies of the number of excess deaths compared to seasonal averages suggest that the reported COVID-19 fatality data also underestimate the true fatality counts associated with COVID-19 [7,8]. This stems in part from the same issue as above: many people who died from the disease at home were never tested and therefore not counted among those confirmed as dying from COVID-19. To correct for this, NYC public health data include both confirmed and probable COVID-related deaths, separately tabulated, and the number of excess deaths suggest that this combination is less incomplete than for reported fatalities in a number of other regions studied. Overall, the excess death counts suggest that reported COVID-19 death counts, particularly those that include probable deaths, underestimate true COVID-related deaths by a much smaller factor than confirmed cases underestimate true infections. Nevertheless, the fatality underestimates mean that the conclusions below about model parameters should be viewed with skepticism; I have made no attempt to correct the fatality data for incompleteness.

I consider fatality data from two sources: (1) the NYC public health site nychealth/coronavirus (data on github), from which I aggregate confirmed plus probable NYC deaths from 3/11/20; (2) the Cook County Medical Examiner site (maps.cookcountyil.gov/medexamcovid19/), with fatality data from 3/16/20. In each case, the first date listed is the date of first fatality attributed by that source to COVID-19, which I denote by *t*1.

From Eqns. (5-7), at early times the cumulative number of deaths is given by

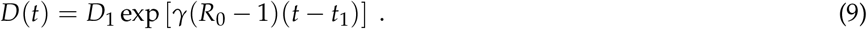

The results of fitting Eqn. (9) to the data from *t*_1_ = 0 to *t*_1_ + 19 days, using the R package `nlstools`, are given in Table 1, with the fits shown in Figs. 1-2. Not surprisingly, the estimates for *D*1 and *γ*(*R*_0_ − 1) are perfectly anti-correlated. Inferred values for *R*_0_ in Table 1 are given for different values of 1/*γ* in the range 4 to 16 days.

**Table 1:** Fits to the first 20 days of fatality data for NYC and Cook County. *t*_1_ is the date of first recorded fatality at that location. *t_d_* is the initial doubling time. Early exponential growth determines the parameter combination *γ*(*R*_0_ − 1) in the 3rd column. Inferred *R*_0_ values in last 3 columns assume *γ*^−1^ = 4, 8, 16 days. Errors on *D*_1_ and *γ*(*R*_0_ − 1) span the 2.5 − 97.5% t-based confidence intervals returned by `nlstools`. These fits are based on minimizing the sum of squares of the residuals between the cumulative data *D*(*t*) and the exponential model. Fits to the daily deaths or that include weighting by Poisson model errors yield values consistent with those in the Table.

| Place | $t_1$ | $D_1$ | $\gamma(R_0 - 1)$<br>(days<sup>-1</sup>) | $t_d$<br>(days) | $\gamma^{-1} = 4$ | $R_0$<br>8 | 16 |
| --- | --- | --- | --- | --- | --- | --- | --- |
| NYC | Mar. 11 | $18.4 \pm 4.8$ | $0.250 \pm 0.014$ | 2.8 | 2.00 | 3.00 | 5.00 |
| Cook County | Mar. 16 | $3.5 \pm 1.1$ | $0.210 \pm 0.017$ | 3.4 | 1.84 | 2.68 | 4.36 |

The selection of 20 days as the interval for the exponential fit for Eqn. (9) is determined by several factors. A significantly shorter time interval would involve smaller data samples and correspondingly noisier parameter estimates. Moreover, I have verified that truncating the fit a few days earlier or starting the fit a few days later does not lead to substantial shifts in the inferred values of *γ*(*R*_0_ − 1). On the other hand, as Figs. 1 and 2 show, extending the fit interval past 20 days would include data impacted by the mitigation measures imposed in mid-March (around time *t_m_*), consistent with the estimate of Δ*t ≃* 20 days for the mean interval from infection to fatality discussed below. This stems from the approximate coincidence in time between the first reported fatalities and the imposition of mitigation measures. Since my interest is in inferring this pre-mitigation parameter combination, I truncate the fit at 20 days.

**Figure 1:**
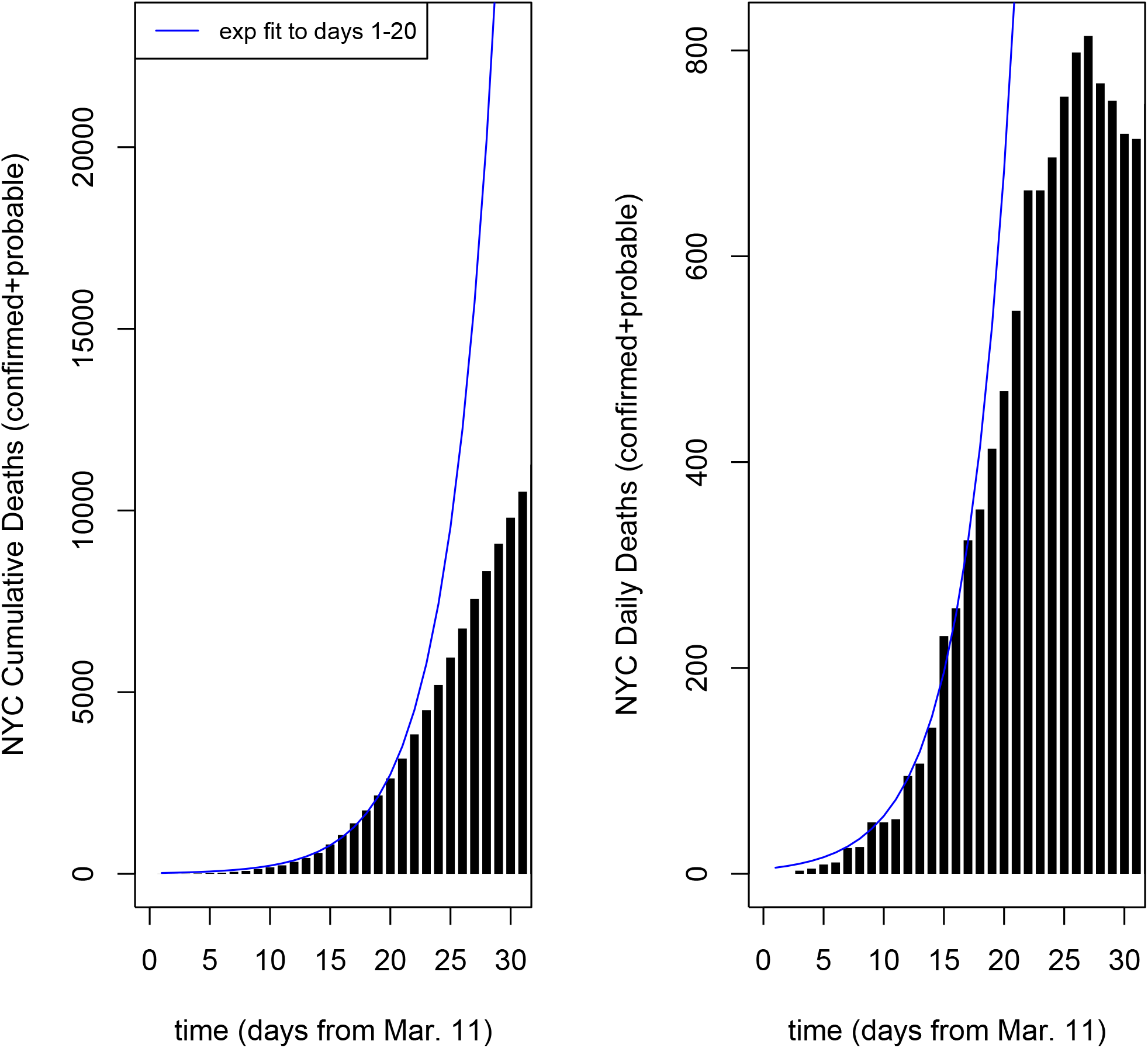
30 days of cumulative (left) and daily (right) COVID-19 related deaths in New York City from the NYC public health site (points). Blue curves show exponential model fit (Eqn. (9)) to cumulative deaths from days 1-20, where day 1 is Mar. 12. Fit parameters are given in Table 1. The impact of mitigation measures taken in mid-March begins to show up as a bend in the data away from the exponential model around the end of March.

**Figure 2:**
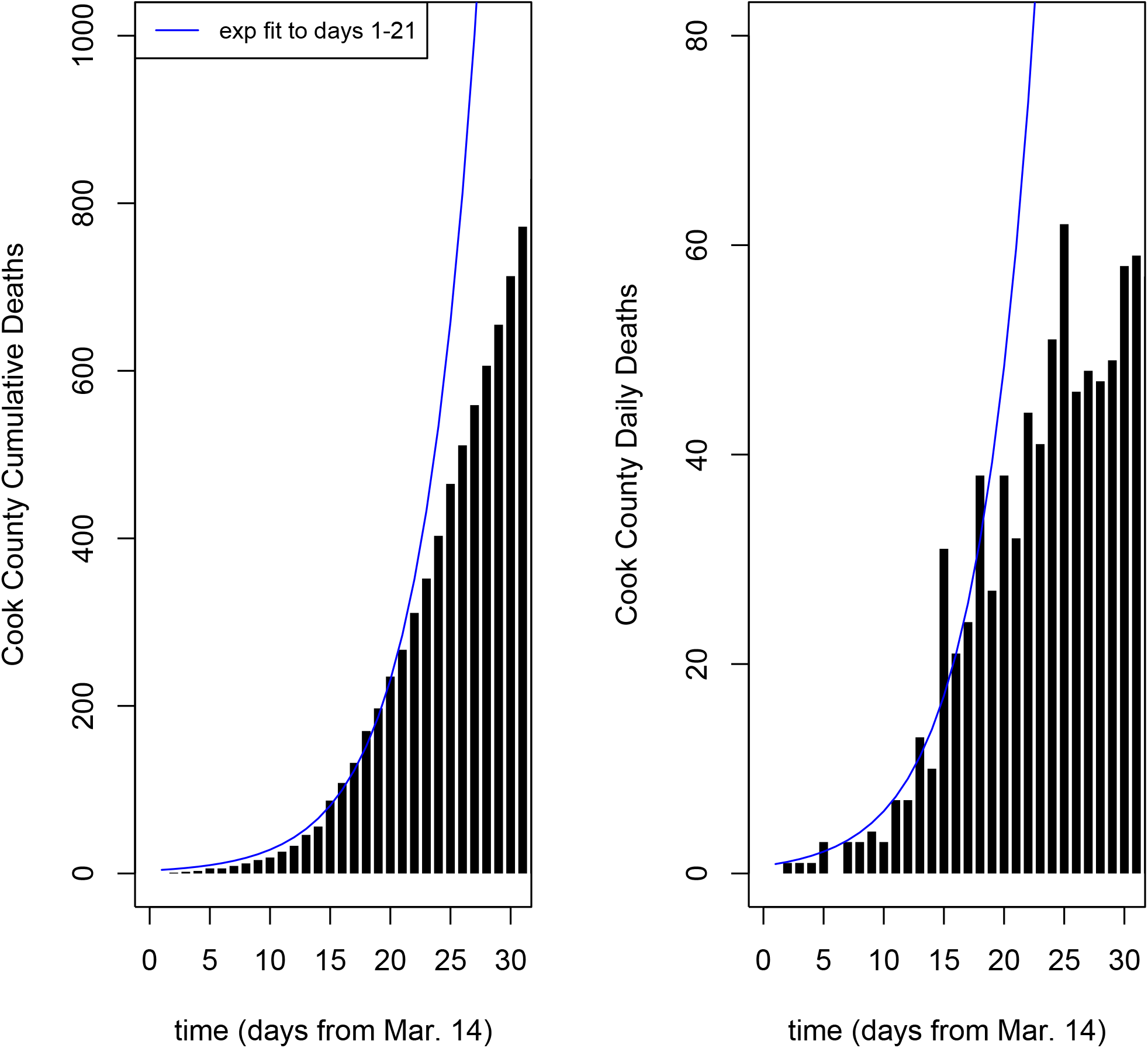
Same as Fig. 1, but for Cook County.

Using the assumed relation between *D*(*t*) and *R*(*t*) and Eqns. (7) and (9), we also have

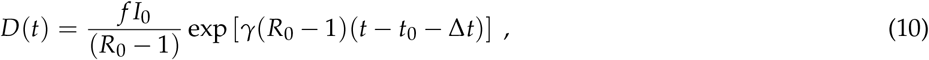

Taking *t*_0_ as the date of the first case(s), for *D*1 ≃ 1 and *I*_0_ ≃ 1, we have

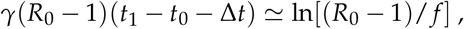

and therefore the time delay between first infections and first deaths is of order *t*_1_ − *t*_0_ ≃ Δ*t* + 20 for *f* = 0.01. If the mean time from infection to death is Δ*t* = 15 − 20 days, then we expect the first deaths to occur of order 35 − 40 days after the first infections. From Table 1, this would put the first infections in early February for both locations. The first *confirmed* cases in both locations were in late February.

## 4 Modeling COVID-19 Evolution

Having fit the early exponential growth of the data, I now explore the evolution of the model and the data over an extended timescale that includes the impact of social contact reduction measures, using the `deSolve` package in R [10] to solve the *SIR* model equations above. In principle, one could carry out a six-dimensional likelihood analysis using Monte Carlo Markov Chain techniques, using either the cumulative fatality data over time, *D*(*t*), or the daily deaths, *dD*/*dt*, to determine credible regions for the parameters *R*_0_, *γ*, *X*, *α*, *t_m_*, and *f* for each city. However, since we have external knowledge of when (*t_m_*) and how rapidly (*α*) mitigation measures were imposed, I first adopted roughly 10-day-wide, flat priors on *t_m_* (centered on mid-March) and on *α*^−1^. I found that the model fits constrain the value of *t_m_* to within less than a day and *α*^−1^ to within 1 − 2 days, so those model parameters are more or less fixed for each location (see below). Given the constraint on *γ*(*R*_0_ − 1) from Table 1, that leaves 3 independent parameters to vary, which I take to be *R*_0_, *X*, and *f*. (One can also treat *γ* as a 4th parameter, using the constraint from Table 1 as a starting value for the non-linear fit, but I find that does not significantly change the results.)

For the results shown below, I take the daily fatality counts as the data vector and assume they can be modeled as an inhomogeneous Poisson process with time-varying intensity, (*dD*/*dt*)_model_ = *λ*(*t*; *R*_0_, *X*, *f*). Given the data, (*dD*/*dt*)*_i_*, on *k* days *t_i_*, one can determine the model parameters by minimizing the negative log-likelihood over the 3-dimensional parameter space,

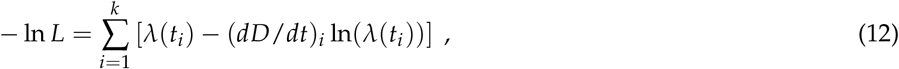

where I have suppressed a term independent of *λ*. In passing from data likelihood to parameter posterior I assume flat priors on the model parameters within a “reasonable” range of variation, which I take to be *f* = 0.1 − 6%, *R*_0_ = 1.1 − 6.0, and *X* = 20 − 99.9%; the flat, bounded prior on *R*_0_ implies a corresponding flat, bounded prior on *γ* via Table 1.

To estimate the goodness-of-fit of the Maximum Likelihood Estimate (MLE) model, one can evaluate the residual deviance for the Poisson likelihood, which is the change in the deviance between the MLE model and the “saturated” model,

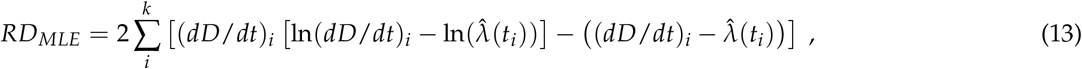

where 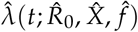 is the MLE model intensity; for *p* model parameters estimated from the data, *RD_MLE_* has a *χ*^2^ distribution with *k* − *p* degrees of freedom (dof) if the model is a good fit. In that case, one can estimate model parameter credible intervals using levels of the *RD* difference, Δ*RD_min_* = *RD*(*R*_0_, *X*, *f*) − *RD_MLE_*.

If the Poisson model errors can be approximated as Gaussian, then with flat priors one could alternatively determine the MLE model parameters and credible ranges by minimizing the usual *χ*^2^ function,

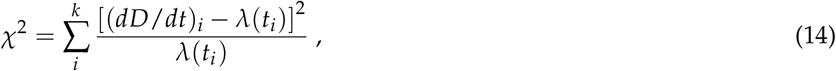

determining goodness-of-fit via the value of 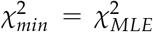 relative to the number of degrees of freedom and parameter intervals via 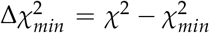 [11]. Since the fatality counts at early and late times are not large compared to unity, it is not obvious at the outset that this would be a good approximation. This is discussed further below.

The extended fatality data alone constrain only a degenerate combination of the 3 parameters; some example model “best-fit” parameters are given in Table 2, with results described in the following subsections.

**Table 2:**
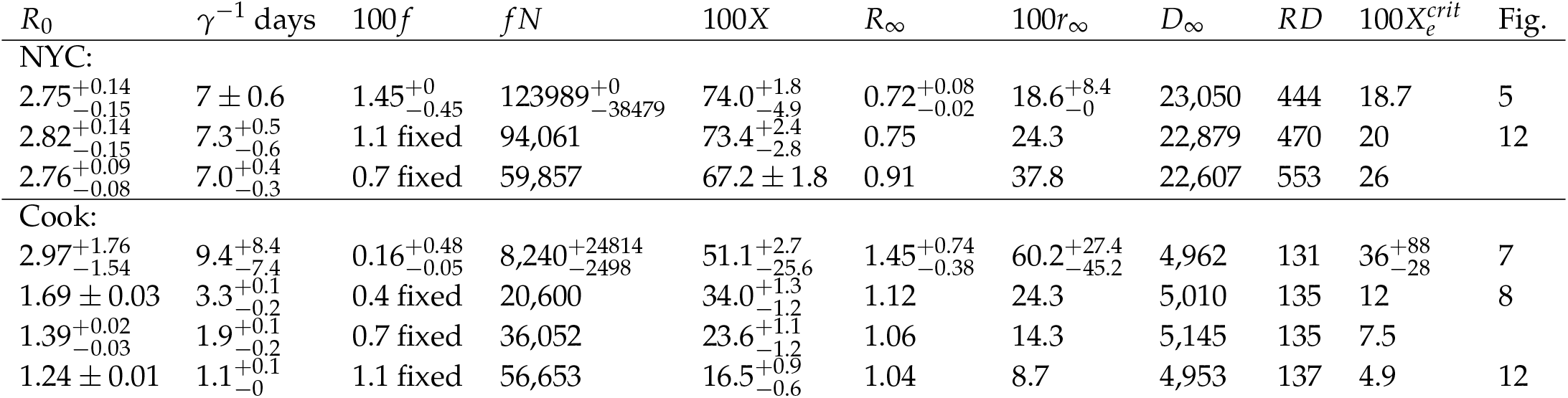
Best-fit *SIR* models with social contact reduction of 100*X*%, with model parameters fit to fatality data for NYC and Cook County, using the tanh mitigation model of Eqn.(8), including estimated 95.45% credible regions. For the first row in each location, credible regions are estimated from simulations of the best-fit model. *R*_0_ and *R*_∞_ are the initial and post-mitigation values of the basic reproduction number, *γ*^−1^ is the mean time to removal from the population after infection, *f* is the infection fatality rate (IFR), *r*_∞_ is the model asymptotic fraction of the population that’s been infected, *D*_∞_ is the model number of asymptotic fatalities, and *RD* is the residual deviance goodness-of-fit measure for the daily fatalities from Eqn. (13); for reference, there are 108 (NYC) and 113 (Cook) degrees of freedom. Due to the perfect degeneracy between *f* and the population *N*, I also indicate the more robust product *fN*, the hypothetical number of asymptotic fatalities if the entire population were infected. Bottom two NYC rows and bottom three Cook County rows indicate fits with IFR *f* fixed by the indicated prior.

| $R_0$ | $\gamma^{-1}$ days | 100*f* | *fN* | 100*X* | $R_\infty$ | 100*r*<sub>∞</sub> | $D_\infty$ | *RD* | 100*X*<sub>*e*</sub><sup>*crit*</sup> | Fig. |
| --- | --- | --- | --- | --- | --- | --- | --- | --- | --- | --- |
| NYC: |  |  |  |  |  |  |  |  |  |  |
| 2.75<sup>+0.14</sup><sub>-0.15</sub> | 7 ± 0.6 | 1.45<sup>+0</sup><sub>-0.45</sub> | 123989<sup>+0</sup><sub>-38479</sub> | 74.0<sup>+1.8</sup><sub>-4.9</sub> | 0.72<sup>+0.08</sup><sub>-0.02</sub> | 18.6<sup>+8.4</sup><sub>-0</sub> | 23,050 | 444 | 18.7 | 5 |
| 2.82<sup>+0.14</sup><sub>-0.15</sub> | 7.3<sup>+0.5</sup><sub>-0.6</sub> | 1.1 fixed | 94,061 | 73.4<sup>+2.4</sup><sub>-2.8</sub> | 0.75 | 24.3 | 22,879 | 470 | 20 | 12 |
| 2.76<sup>+0.09</sup><sub>-0.08</sub> | 7.0<sup>+0.4</sup><sub>-0.3</sub> | 0.7 fixed | 59,857 | 67.2 ± 1.8 | 0.91 | 37.8 | 22,607 | 553 | 26 |  |
| Cook: |  |  |  |  |  |  |  |  |  |  |
| 2.97<sup>+1.76</sup><sub>-1.54</sub> | 9.4<sup>+8.4</sup><sub>-7.4</sub> | 0.16<sup>+0.48</sup><sub>-0.05</sub> | 8,240<sup>+24814</sup><sub>-2498</sub> | 51.1<sup>+2.7</sup><sub>-25.6</sub> | 1.45<sup>+0.74</sup><sub>-0.38</sub> | 60.2<sup>+27.4</sup><sub>-45.2</sub> | 4,962 | 131 | 36<sup>+88</sup><sub>-28</sub> | 7 |
| 1.69 ± 0.03 | 3.3<sup>+0.1</sup><sub>-0.2</sub> | 0.4 fixed | 20,600 | 34.0<sup>+1.3</sup><sub>-1.2</sub> | 1.12 | 24.3 | 5,010 | 135 | 12 | 8 |
| 1.39<sup>+0.02</sup><sub>-0.03</sub> | 1.9<sup>+0.1</sup><sub>-0.2</sub> | 0.7 fixed | 36,052 | 23.6<sup>+1.1</sup><sub>-1.2</sub> | 1.06 | 14.3 | 5,145 | 135 | 7.5 |  |
| 1.24 ± 0.01 | 1.1<sup>+0.1</sup><sub>-0</sub> | 1.1 fixed | 56,653 | 16.5<sup>+0.9</sup><sub>-0.6</sub> | 1.04 | 8.7 | 4,953 | 137 | 4.9 | 12 |

An important timescale for SIR inference from the fatality rate is the mean time from infection to death; the early study of [3] suggests it is of order 15 − 20 days, which provides an estimate of the time-delay parameter Δ*t* introduced above. I adopt 20 days as the fiducial value for Δ*t*; changing this value has no direct impact on the fatality-based modeling and just shifts the model curves for *s*, *i* and *r* in time. Equivalently, fixing Δ*t* translates the mitigation time parameter *t_m_* to a date in real time. As we shall see, the resulting dates for *t_m_* are consistent with the mid-March time frame when contact reduction measures were implemented in both locations, indicating that this choice of Δ*t* is consistent with that external information.

### 4.1 New York City

For the NYC model fits, I adopt the tanh mitigation model of Eqn.(8) and take the NYC population to be *N* = 8, 550, 971 (as noted above, *N* and *f* can be relatively rescaled, keeping the product fixed, to yield identical results). I use the daily NYC public health fatality data vs. time, including confirmed plus probable deaths, through June 30, 2020 (day 111), as downloaded on July 2. As of that date, there had been 23,059 cumulative NYC fatalities reported.

Using fatality data alone, the Poisson model MLE parameters are *R*_0_ = 5.15, *X* = 99.2%, *f* = 4.9%, with *t_m_* = *t*_1_ + 22.9 − Δ*t* = Mar. 14 and *α*^−1^ = 7.7 days as the time interval over which mitigation measures were implemented; the data vs. model comparison is shown in Fig. 3. For these model parameters, the June 30 cumulative fatality total is 23,126 (within 0.3% of the actual value), the asymptotic value is *D*_∞_ = 23, 256, the asymptotic value of *R_t_* is *R*_∞_ = 0.04, and 100*r*_∞_ = 5.6% of the population, or 470,596 people, will end up having been infected. The dashed vertical line in the upper left panel of Fig. 3 indicates the date when *Rts* = 1, which is when the peak in the number of model infected occurs, around Mar. 18 in this example; on that date, in the model 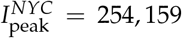 people in NYC had the infection. The residual deviance is *RD* = 401.5 for *k* − p = 108 degrees of freedom; while this is quantitatively a poor fit, the data-model comparison in Fig. 3 indicates that the model appears to capture the key qualitative behaviors and detailed shape of the data; for more discussion, see §5. The fatality predictions to June 30 are in principle sensitive to the model curves to June 30 −Δ*t* = June 10, or about Day 93. In the MLE model, the initial serial time is 〈*t_s_*〉 = 1/*R*_0_*γ* = 3.2 days, and the initial removal time is *γ*^−1^ = 16.6 days. For these model parameters, as Fig. 3 shows, for *t* ≿30 days, *R_t_ ~* 0, so there were relatively few new infections after mid-April, and the number of infected thereafter decayed exponentially, *I* ~ *I*_30_ exp[−*γ*(*t* − 30)]; similarly, (*dD*/*dt*)(*t*) ~ (*dD*/*dt*)_50_ exp[−*γ*(*t* − 50)]. Since *R*_0_ > 2, the exponential decline was slower than the initial exponential growth.

**Figure 3:**
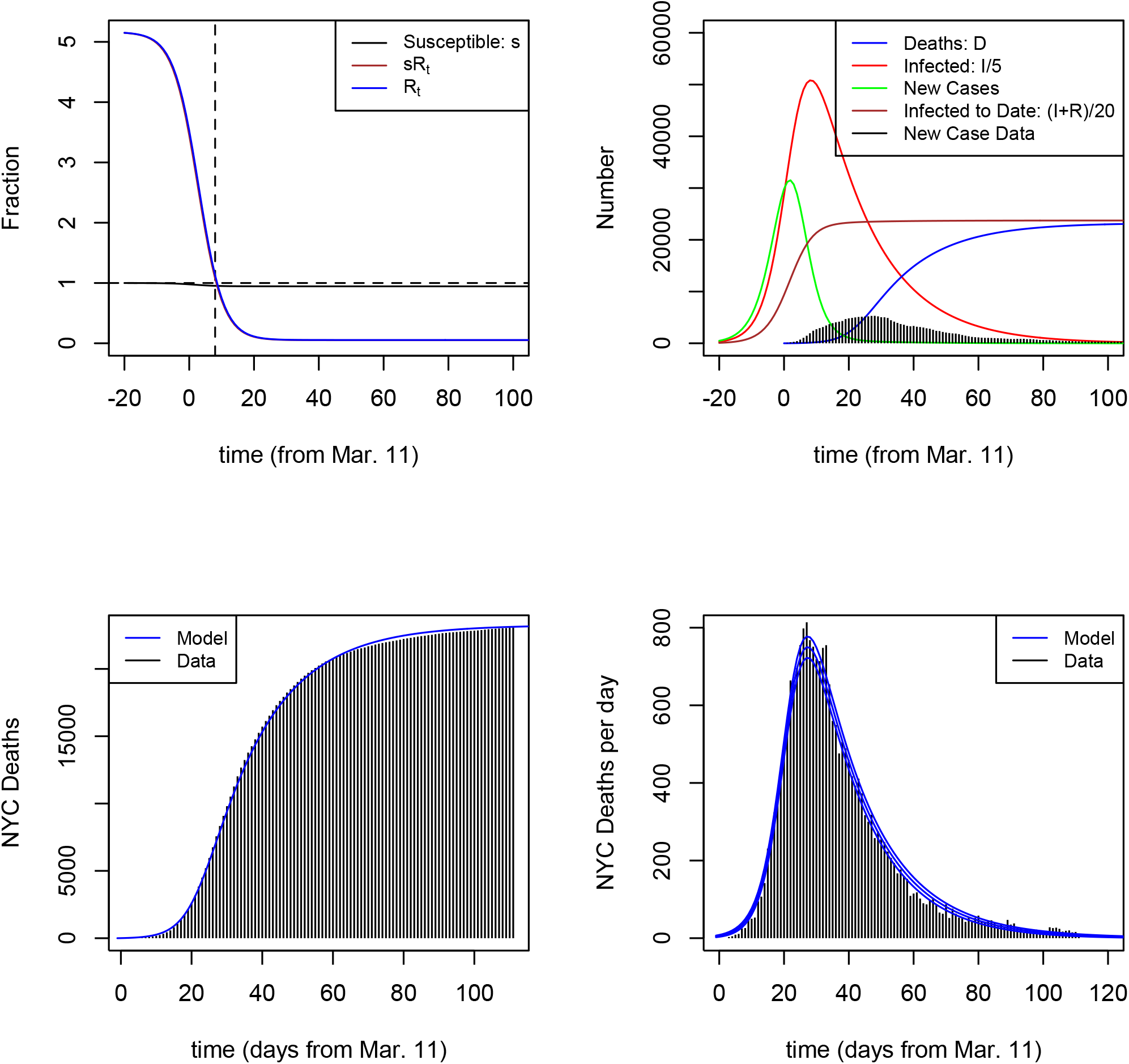
Fatality-only MLE SIR model for NYC, with *R*_0_ = 5.15 and 100*X* = 99.2% reduction in social contact implemented over an 8-day period in mid-March, with IFR *f* = 4.9%. Model parameters were fit using daily fatality data *dD*/*dt* through June 30. *Upper left:* Fraction of susceptible, *s* (black), parameter *R_t_* (blue), and product *R_t_s* (brown) vs. time. *Upper right:* Number infected at a given time divided by 5, *I*/5 (red), new infections per day (green), cumulative number infected divided by 20 (brown), cumulative number of deaths (blue), and reported case data (histogram). *Lower left:* cumulative number of reported deaths (histogram) and model (blue curve). *Lower right:* daily fatalities, data (histogram) and model (blue curve); upper and lower curves indicate ±1 − *σ* intervals, assuming model daily deaths are Poisson-distributed.

If the MLE model were a good fit, then we could determine parameter uncertainties from where the value of Δ*RD_min_* = *RD*(*X*, *R*_0_, *f*) − *RD_MLE_* is below 1, 4 and 9; these regions would be expected to delimit 68, 95.5, and 99.7% credible limits for one parameter. Under this assumption, I find 95.45 (99.7)% ranges of *R*_0_ = 3.56 − 5.34(3.15 − 5.38), *X* = 87.8 − 99.8(83.0 − 99.8)%, and *f* = 3.1 − 5.9(2.7 − 5.9)%. Note that the upper bounds on *X* and *R*_0_ come from imposing the physical constraint that *X* < 1, while the upper bound on *f* is just the upper bound on the imposed prior, so the data do not constrain these model parameters at the upper end. The parameters *R*_0_ and *X* are highly correlated: larger values of *R*_0_ require greater degrees of social contact reduction, *X*, in order to match the data. The degeneracy is well-approximated by the expression *X* − 1 ≃ 0.24 − (1.28/*R*_0_), which implies *R*_∞_ ≃ 1.28 − 0.24*R*_0_, that is, larger *R*_0_ requires smaller *R*_∞_ to fit the NYC fatality data. The requirement *X* < 1 or equivalently *R*_∞_ > 0 also implies the upper bound *R*_0_: 5.33.

#### 4.1.1 Inclusion of Case Data

The preceding analysis constrained *SIR* parameters for NYC using only fatality data. However, the upper right panel of Fig. 3 shows that the MLE model for the fatality data underpredicts the reported NYC case data by a significant margin (green curve vs. histogram). Here, I introduce constraints from NYC public health data on reported daily COVID-19 cases, (*dC*/*dt*)*_d_*. Using Eqn. (21) from Appendix B, we can write the model daily rate of new cases as

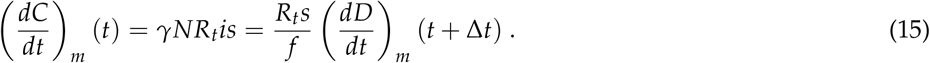

Since the reported case data provides a lower bound on the true number of cases (not every infected person gets tested or tests positive and also assuming that the false positive rate is low), the model prediction for *dC*/*dt* must exceed the data value, i.e., (*dC*/*dt*)*_m_* ≥ (*dC*/*dt*)*_d_*, which implies

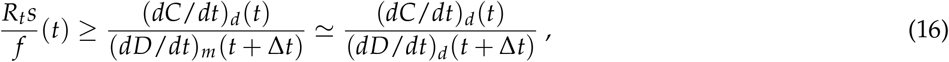

where the last equality holds for models that approximately reproduce the daily fatality data. An example of this is shown in the left panel of Fig. 4, which plots the far left and right hand sides of Eqn. (16). The RHS (points) comes from the daily case and fatality data from the NYC public health site, using Δ*t* = 20 days, while the LHS is the blue curve for the best-fit model satisfying the case bound, discussed below.

**Figure 4:**
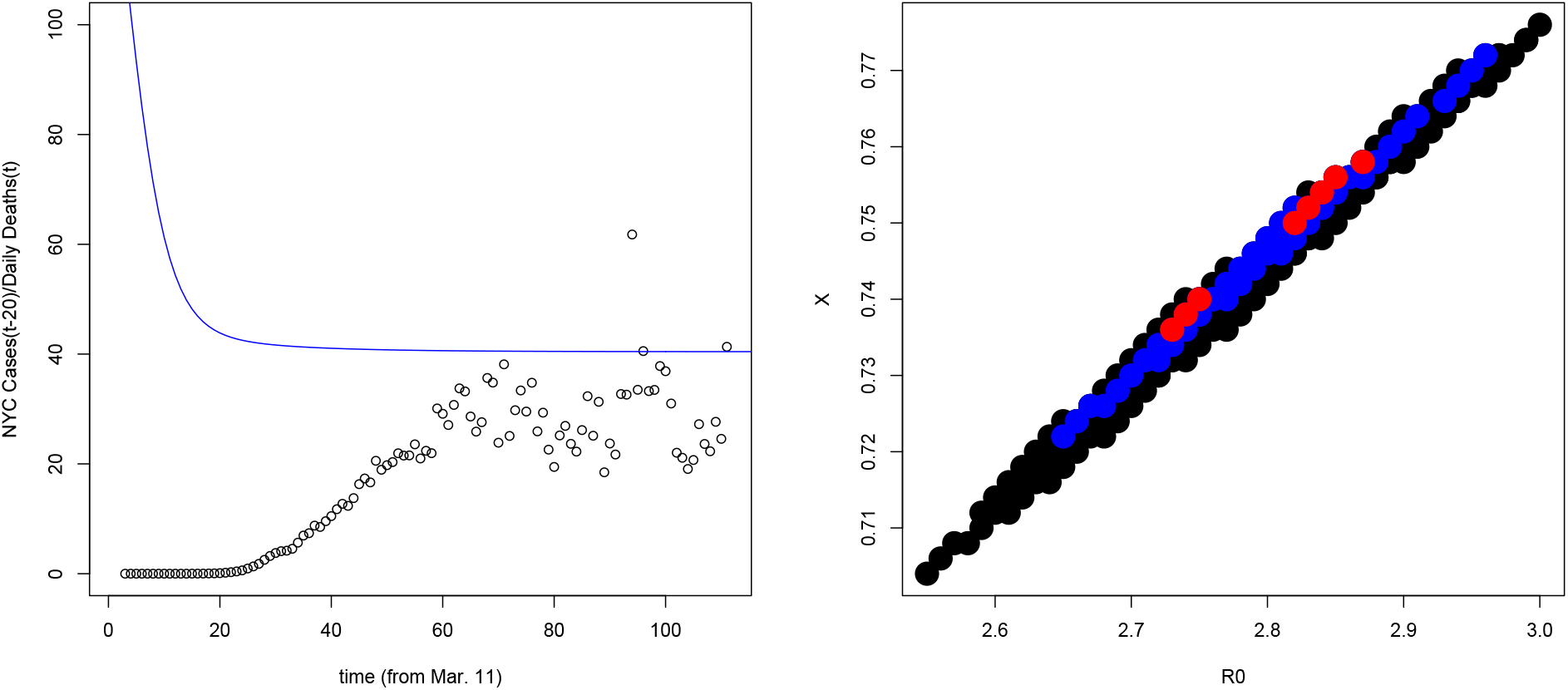
*Left:* Ratio of time-shifted, 7-day average of NYC new case data to fatality data, (*dC*/*dt*)*d*(*t* − 20)/(*dD*/*dt*)*_d_*(*t*), the RHS of Eqn. (16) (points). Blue curve shows *R_t_s*/ *f* for the best-fit NYC model with model cases greater than reported cases, with *R*_0_ = 2.75, *X* = 74%, and *f* = 1.45% (model of Fig. 5). If the case data were complete, it would track the model curve for the true model. *Right:* Degeneracy in the *R*_0_ vs. *X* plane of models that satisfy the case data bound of Eqn. (16). Red, blue, and black points have Δ*RD* = *RD* − *RD_min_* < 1, 4, 9 respectively, relative to the model of Fig. 5.

The left panel of Fig. 4 indicates that, not surprisingly, cases were undercounted by a very large factor at early times but then ramped up linearly as testing became more widespread. By day 60 − Δ*t* = 40, or about April 20, the reported case rate reached an approximate plateau as a fraction of the future fatality rate and thus, according to the *SIR* models, as a fraction of the true case rate.

At late times, the LHS of Eqn. (16) is well approximated by *R*_0_(1 − *X*)(1 − *r*_∞_)/ *f* = *R*_0_(1 − *X*)(1 − *D*_∞_/ *fN*)/ *f*, and the left panel of Fig. 4 indicates that this combination of model parameters should be bounded below by about 40 in order to satisfy the case data lower bound. Imposing this as a sharp prior in the 3-dimensional parameter space (and setting *D*_∞_ = 23,000 in the above expression) leads to the upper bound *f* ≤ 1.75%. The best-fit model satisfying this constraint has *R*_0_ = 2.75, *X* = 74% (and thus *R*_∞_ = 0.72), *γ*^−1^ = 7 days, and *f* = 1.45%, with *RD_min_* = 444, *r*_∞_ = 18.6%, *D*_∞_ = 23,050, *D* = 22,932 on June 30, and initial serial time 〈*t_s_*〉 = 1/*R*_0_*γ* = 2.5 days; see Fig. 5 and first row of Table 2. For these parameter values, the case data lower bound is saturated, that is, the reported daily cases from April 20 onward were a nearly complete tally of the true cases; this is shown in the left panel of Fig. 4.

**Figure 5:**
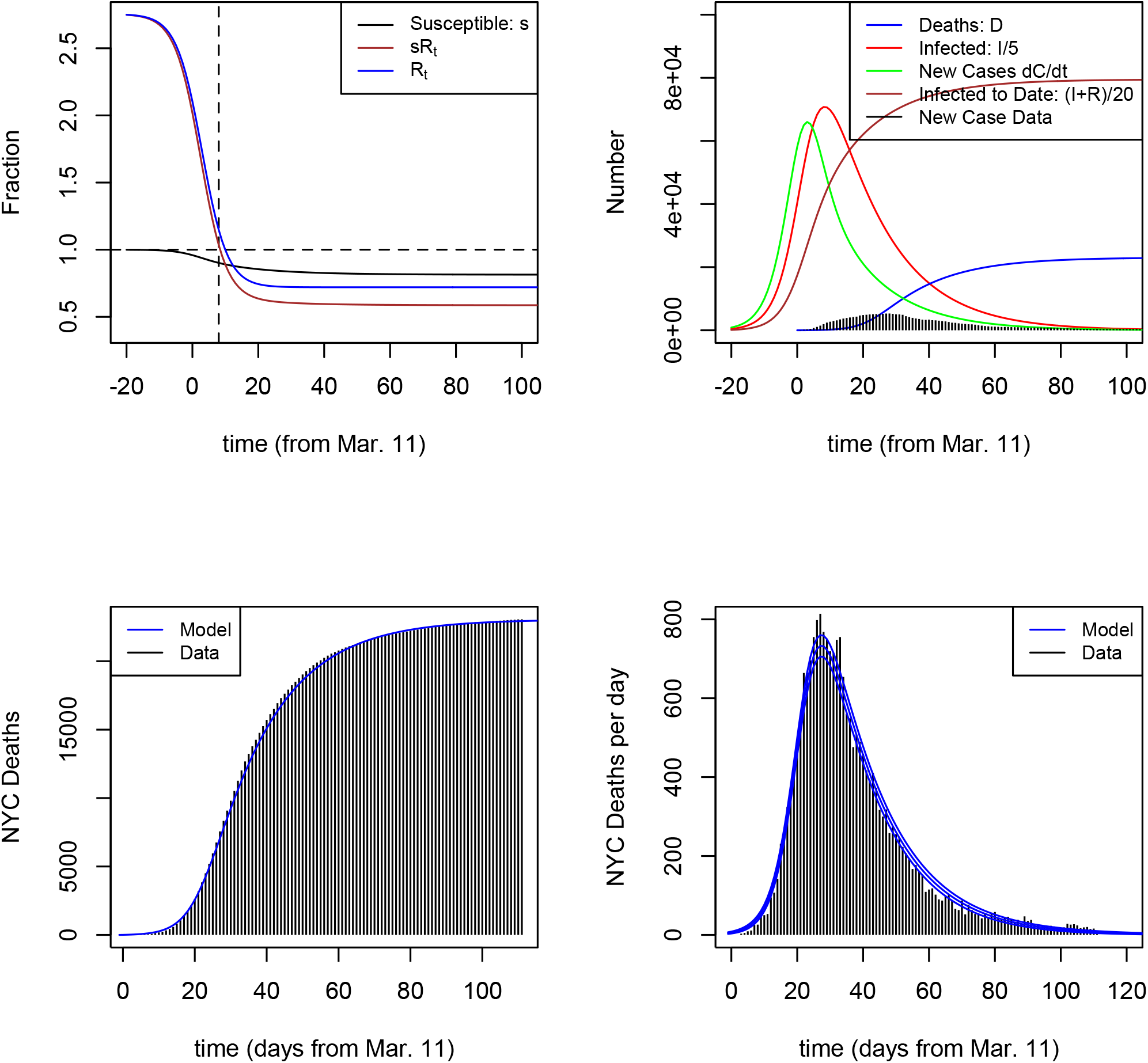
Similar to Fig. 3, but for a model with *R*_0_ = 2.75, *X* = 74%, *f* = 1.45%, which is the best-fit model that satisfies the case data lower bound, as shown in the upper right panel. *RD* = 444 for this model. In this case, *γ*^−1^ = 7 days, *R*_∞_ = 0.72, asymptotic model NYC death total is *D*_∞_ = 23,050, and *r*_∞_ = 18.6% of the population get infected.

The *R*_0_ vs. *X* degeneracy in this case-restricted parameter space is shown in the right panel of Fig. 4; using Δ*RD* = *RD* − *RD_min_* < 9, which would correspond to the 99.7% credible range if the best-fit model were a good fit, we find *R*_0_ = 2.55 − 3.00, *X* = 70.4 − 77.6%, and *f* = 1.30 − 1.55%. Simulations provide another approach to parameter uncertainty estimation. Fig. 6 shows the MLE parameter estimates inferred for 1000 simulations of the best-fit model (Fig. 5), where each simulation is a Poisson sample drawn from the model intensity, *λ_min_*(*t*). For computational speed, the fit for each simulation is done by minimizing the *χ*^2^ statistic of Eqn. (14); the lower right panel shows that *χ*^2^ and *RD* are tightly correlated, so this procedure is effectively identical to minimizing *RD* for each simulation. The red points show those parameter values that also satisfy the case data constraint of Eqn. (16); note that the model being simulated lies on the boundary of the red region. From the red points, the inferred 95.45% (99.7%) credible ranges from the simulations are: *R*_0_ = 2.60 − 2.89(2.54 − 2.98), *X* = 69.1 − 75.8%(66.9 − 77.2%), *f* = 1.0 − 1.43%(0.88 − 1.50%), in reasonable but not perfect agreement with the Δ*RD* estimates from the model fits to the data; I use the simulation-based intervals in Table 2.

**Figure 6:**
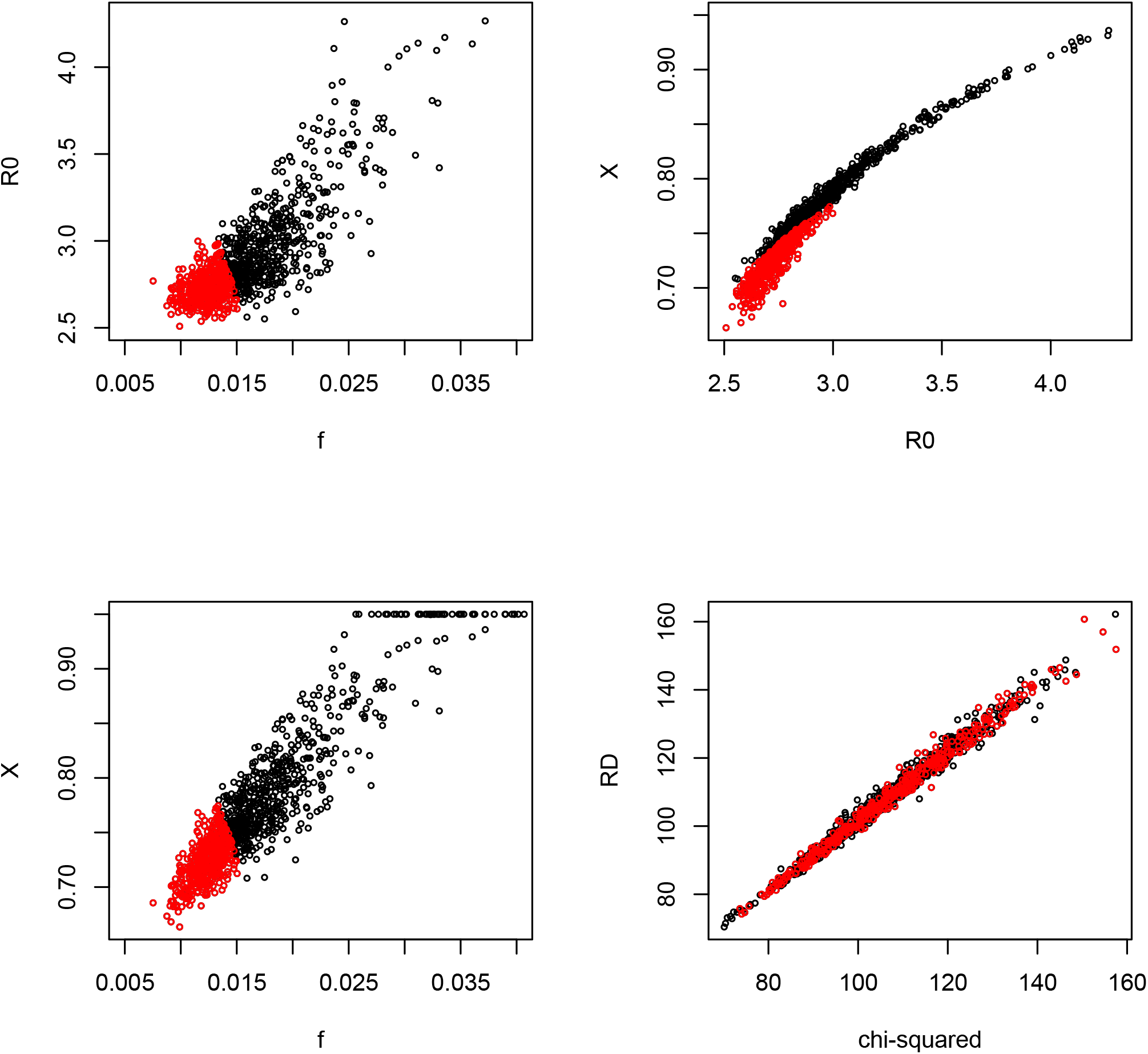
Results from 1000 simulations of the best-fit NYC *SIR* model that satisfies the case data lower bound (see Fig. 5). Each point corresponds to the best-fit in the two-dimensional subspace for that simulation, using the *χ*^2^ statistic of Eqn. (14). *Upper left: f* vs. *R*_0_; *Upper right: R*_0_ vs. *X*; *Lower left: f* vs. *X*; *Lower right: RD* vs. *χ*^2^ statistic. Red points correspond to model parameters that satisfy the case data lower bound of Eqn. (16).

The models above suggest a preference for relatively high values of the NYC IFR, *f* > 1%. If there were a strong external constraint (prior) on *f* from other studies that indicated lower values, the preferred values for the other model parameters would shift accordingly. To demonstrate this, the 2nd and 3rd rows of Table 2 indicate the best-fit models to the daily fatality data with imposed priors of *f* = 1.1 and 0.7%. Table 2 suggests the value of *R*_0_ ≃ 2.8 is well-determined, independently of *f*, for NYC.

A feature of the mitigation models is that the asymptotic value of *r*_∞_ for models that fit the fatality data depends on the Infection Fatality Rate *f* as roughly 1/ *f*. This is expected since *D*_∞_ = *Nfr*_∞_, so *r*_∞_ = *D*_∞_/*Nf*, and for NYC the model prediction for *D*_∞_ is well-constrained by the data. This inverse trend of *r*_∞_ with *f* is evident in Table 2. This scaling also enables us to place a very conservative lower bound on *f*, since it implies *f* = *D*_∞_/*Nr*_∞_ > *D*_∞_/*N* > *D*(June 30)/*N* = 0.27% for NYC.

#### 4.1.2 Predictivity of the NYC Model

The foregoing analysis shows that the simple *SIR* model with a single transition in social contact appears to qualitatively describe the NYC fatality data through the end of June, 2020. In fact, by examining earlier subsets of the data, one can infer that the model became *quasi-predictive* relatively soon after the peak in fatalities, in the sense that the number of subsequent fatalities over a significant time interval were accurately predicted. As an example, using fits to fatality data through April 22 (Day 43), at which time there were 15,864 NYC confirmed plus probable fatalities in the public database, the best-fit models predicted the subsequent month’s 5,151 fatalities (April 23 to May 20) to about 1.3% accuracy, about equal to the statistical error. The success of this prediction is expected to be limited in part by the fact that NYC continually updates its fatality data for earlier dates, sometimes with significant additions for dates weeks earlier; e.g., by July 2, the cumulative reported fatalities through April 22 had increased from 15,864 to 16,129.

### 4.2 Cook County

For the Cook County model fits, I take the Cook County population to be *N* = 5, 150, 233. I use public health fatality data through July 9 (day 116), downloaded from the Cook County Medical Examiner site on July 12; as of July 9, total reported fatalities were 4,725.

Fig. 7 shows the MLE tanh *R_t_* model fit to the daily fatality data, with *R*_0_ = 2.97, *X* = 51.1%, *f* = 0.16%, *t_m_* = *t*_1_ + 20.6 − Δ*t* = Mar. 17 (2 days earlier relative to the first fatality than for NYC), and *α*^−1^ = 3.3 days (4 days shorter than for NYC); asymptotically, 100*r*_∞_ = 60.2% of the population get infected, and *D*_∞_ = 4, 962. The goodness-of-fit *RD* = 131 for 113 degrees of freedom, which is acceptable. The model prediction for July 9 was *D* = 4, 746 fatalities, within 0.5% of the data value and well within the expected statistical error. The initial serial time in the Cook County MLE model is 〈*t_s_*〉 = 3.2 days. At the peak, 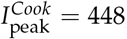, 109 individuals were infected in the MLE model. The feature in the curve of new infections vs. time (green curve in upper right panel) is due to the sharply falling behavior of *R_t_* around that time.

**Figure 7:**
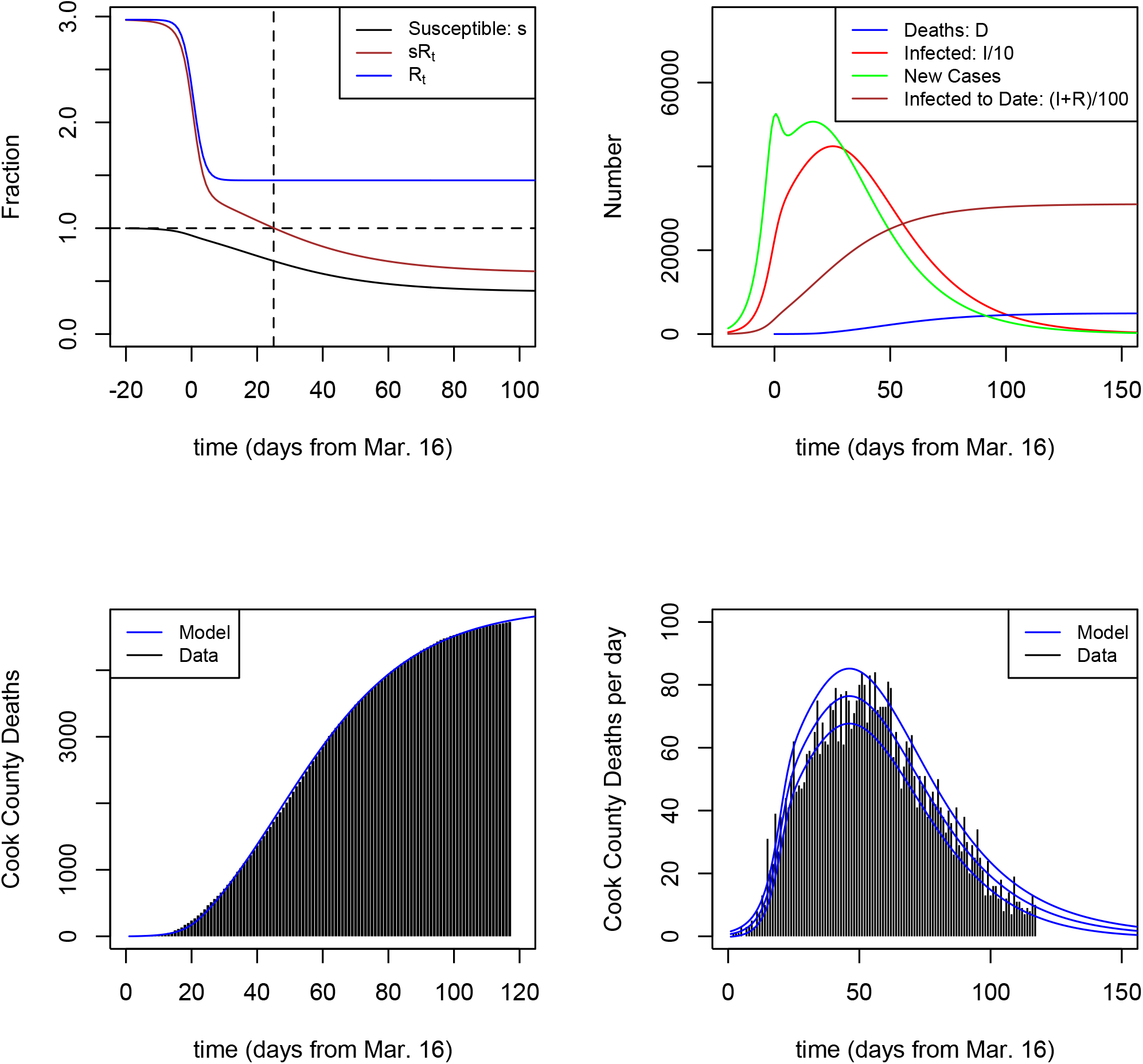
SIR MLE model for Cook County daily fatality data through July 9, with *R*_0_ = 2.97, *X* = 51.1%, and *f* = 0.16%, with measures implemented over an *α*^−1^ = 3.3-day period. Upper and lower model curves in lower right panel indicate ±1 − *σ* intervals, assuming model daily deaths are Poisson-distributed, suggesting that the day to day variations in Cook County reported fatalities are roughly consistent with statistical fluctuations.

As another example, Fig. 8 shows the best-fit model with a sharp prior of *f* = 0.4%, with *R*_0_ = 1.69 and *X* = 34%, which has a goodness-of-fit parameter *RD* = 135 for 114 degrees of freedom (Δ*RD_min_* = 4). The model prediction for July 9 was *D* = 4, 786, which agrees with the data to 1.2%. In this model, asymptotically 100*r*_∞_ = 24.2% of the population will be infected, and *D*_∞_ ≃ 4, 995 fatalities. The 3rd and 4th Cook County entries in Table 2 show best-fit results for two other values of *f*, 0.7 and 1.1%.

**Figure 8:**
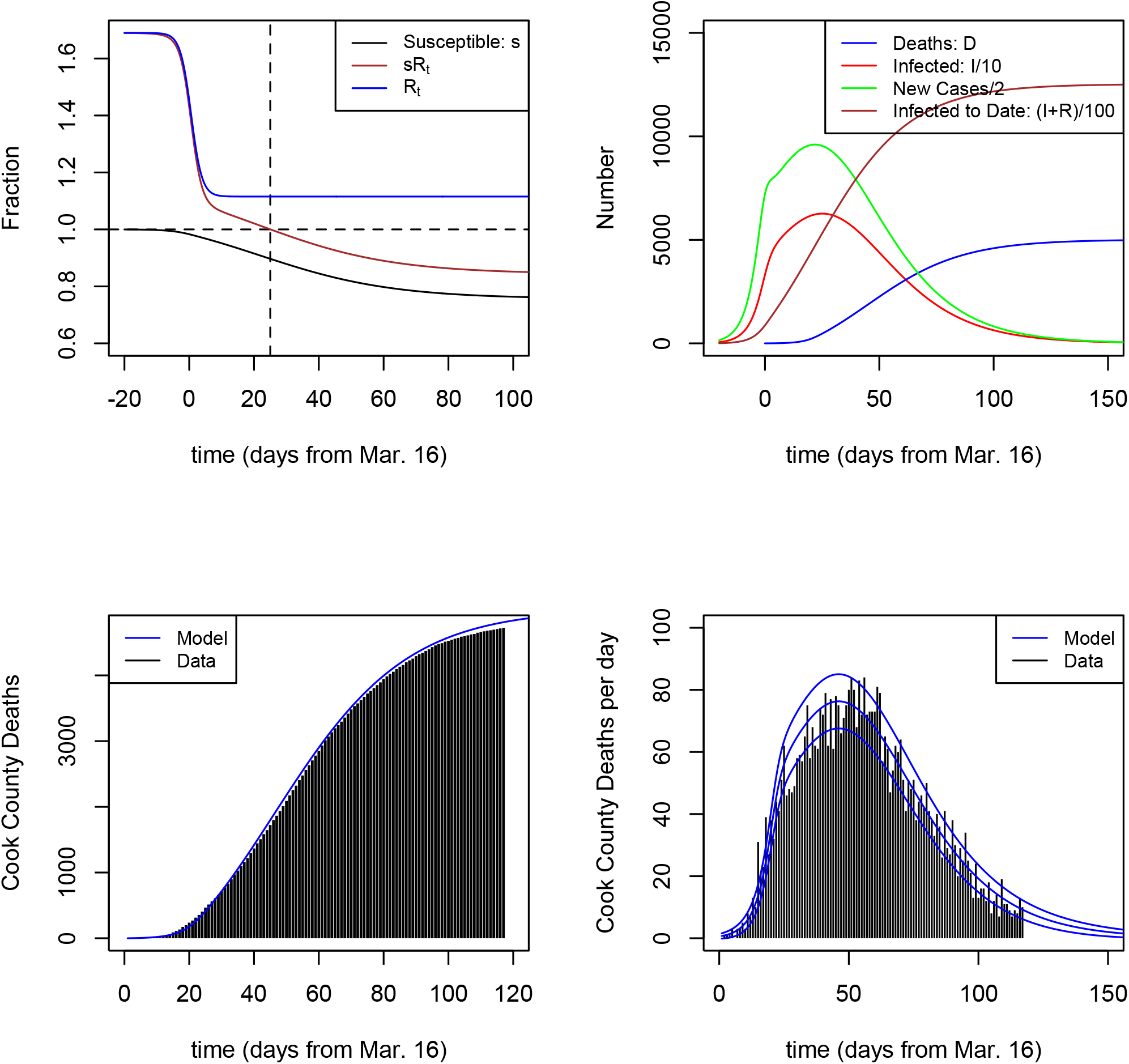
SIR model for Cook County data through July 9, showing best-fit model for daily fatalities with fixed prior of *f* = 0.004, for which *R*_0_ = 1.69, *X* = 34%, with measures implemented over an *α*^−1^ = 3.3-day period (2nd Cook County row in Table 2).

The striking feature of the Cook County data, compared to NYC, is the roughly linear growth in cumulative fatalities over an extended time period from day ≃ 35 − 70 and a corresponding extended plateau in daily fatalities. Since the daily fatality rate *dD*/*dt* = *f γI*, linear growth in *D* is achieved if *I* ≃ *I_peak_* = constant over an extended time period, which requires *R_t_s ≃* 1 and very slowly varying. Since both *R_t_* and *s* are monotonically decreasing, this requires *R_t_* ≃ 1 + *δ*, *s* ≃ 1 − *δ*, in order to achieve this slower evolution. Indeed, Table 2 shows that for the 2 models shown here, the asymptotic value of *R*_∞_ is modestly above unity. This behavior is analogous to the phenomenon of “critical slowing down” in the approach to equilibrium in second-order phase transitions and in dynamical systems, which also involve a transition from exponential to power-law evolution—for discussion, see Appendix A. From mid-April through late May, in Cook County *dD*/*dt* ≃; 70 deaths per day; this implies 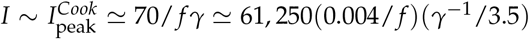 were infected at that time, with a comparable number of new cases per day, as shown in Figs. 7 and 8.

The model parameter degeneracy for Cook County is shown in Fig. 9. Using Δ*RD* = 4, 9, the approximate 95.45 (99.7)% credible intervals are *R*_0_ = 1.39 − 4.18(1.21 − 4.85), *X* = 23.6 − 54.5%(14.8 − 55.4%), and *f* = 0.12 − 0.7%(0.11 − 1.5%). The locii are roughly approximated by *R*_0_ ≃ 1 +(0.003/ *f*) (for *f* > 0.002) and *R*_0_ ≃ 0.005/ *f* (for *f* < 0.002), and by the not very illuminating polynomials 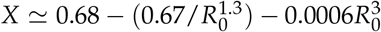 and *X* ≃ 0.65 − 80 *f* (for *f* < 0.004) and *X* ≃ 0.55(0.002/ *f*)^0.7^ (for *f* > 0.004).

**Figure 9:**
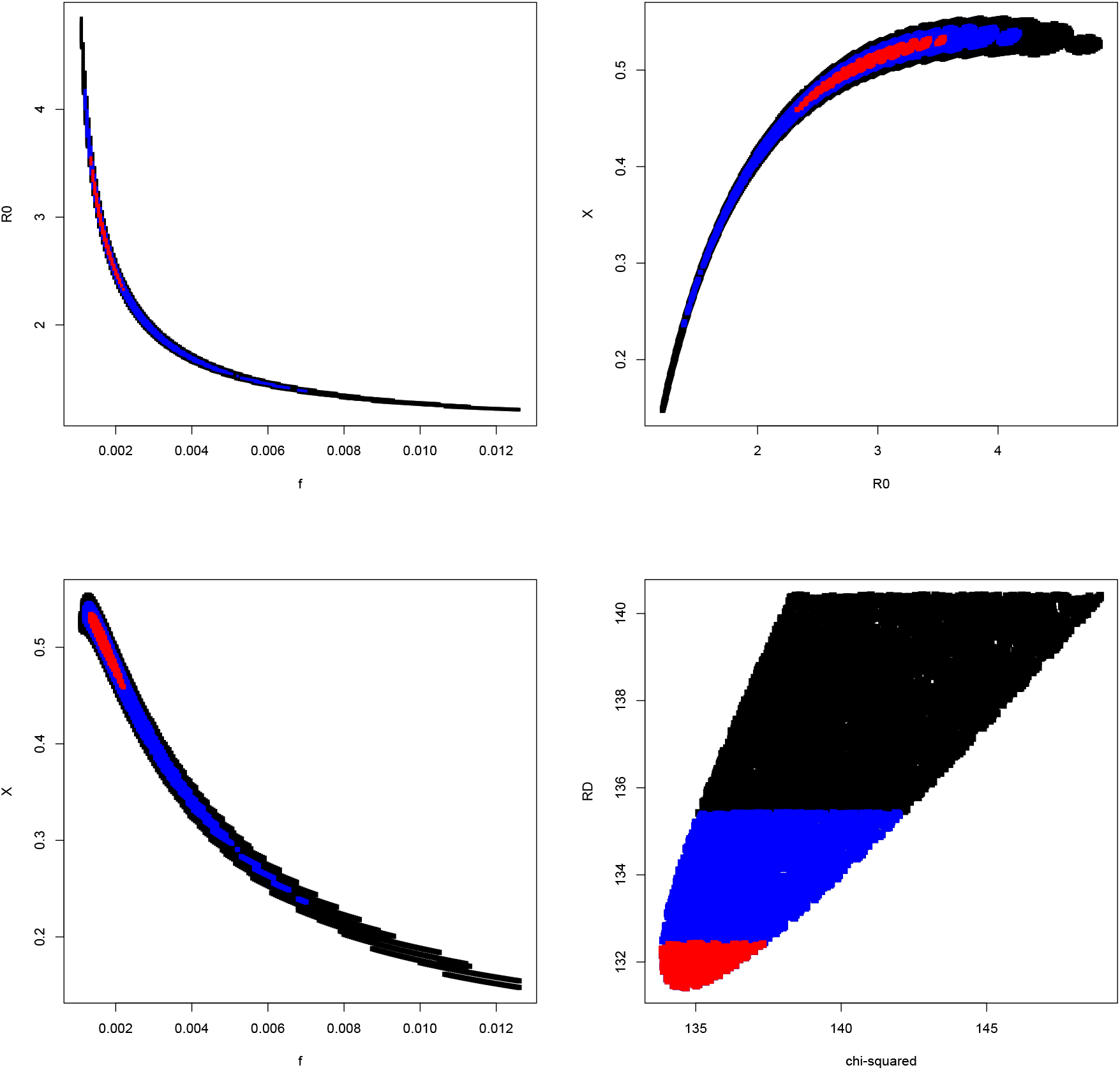
Distribution of minimum values of Δ*RD_min_* = *RD*(*X*, *R*_0_, *f*) − *RD_MLE_* in the two-dimensional parameter subspaces for Cook County, projected over the third parameter. Red, blue, and black points indicate models with Δ*RD_min_* < 1, 4, 9, which would correspond to 68, 95.5, and 99.7% credible limits for one parameter if the MLE model were a good fit. *Upper left: f* vs. *R*_0_; *Upper right: R*_0_ vs. *X*; *Lower left: f* vs. *X*; *Lower right: RD* vs. *χ*^2^ statistic.

As for NYC, we can alternatively estimate parameter uncertainties via simulation. Fig. 10 shows the results of model fitting 500 simulations of the Cook County MLE model. The simulated parameter degeneracies are close to those inferred from the data-model fit, and we find simulated 95.45% credible intervals of *R*_0_ = 1.43 − 4.73, *X* = 25.5 − 53.8%, and *f* = 0.11 − 0.64%, comparable to the Δ*RD* = 4 results above, while the expected (4.55 − 95.45)% range for the goodness-of-fit parameter is *RD* = 91.7 − 140.8, which encompasses the MLE value for the data. As for NYC, I adopt the simulation-based intervals in Table 2.

**Figure 10:**
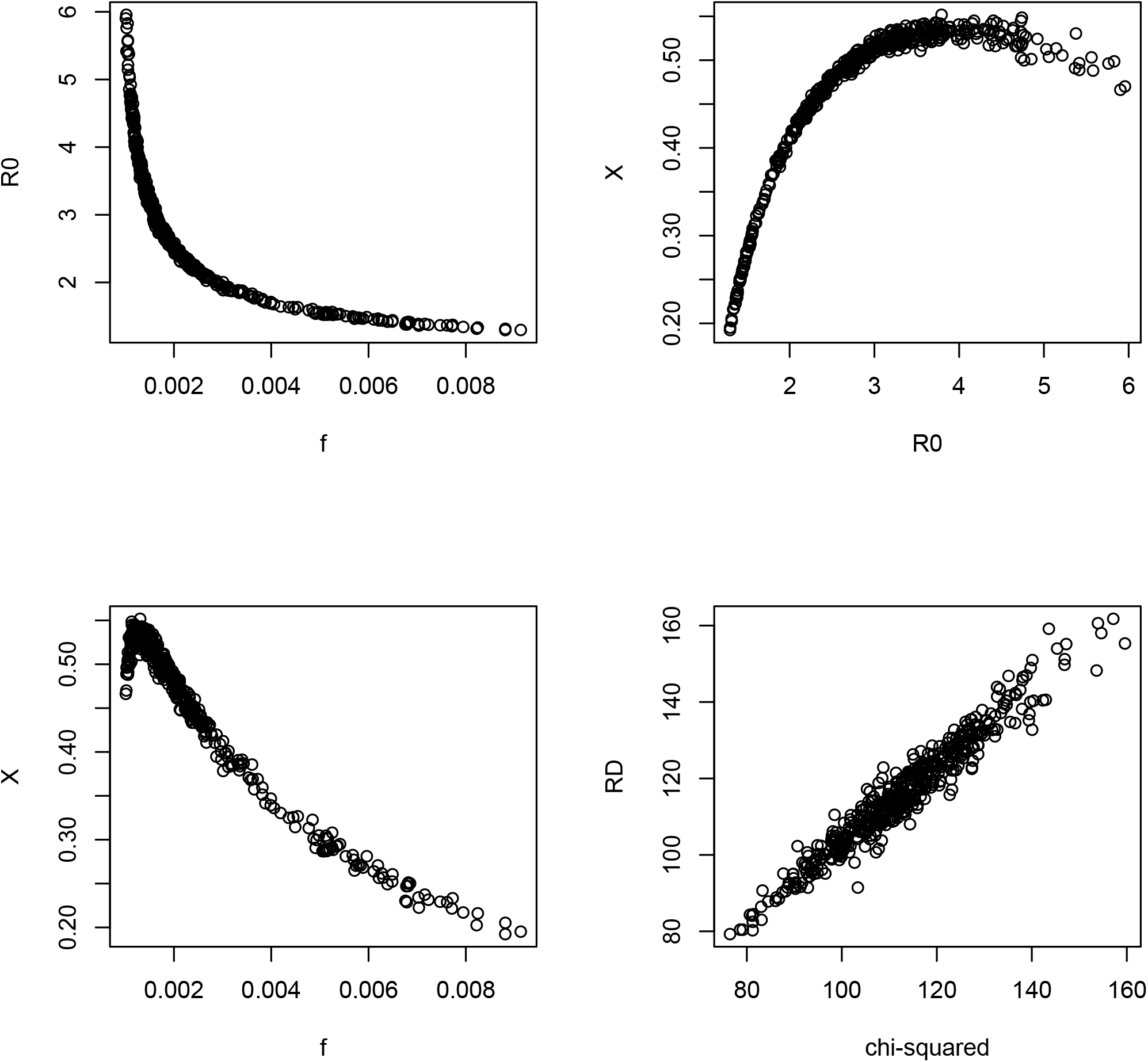
Same as Fig. 9 but now for 500 simulations of the Cook MLE model, showing best-fit models for each simulation using the *χ*^2^ statistic of Eqn. (14).

For Cook County, we can draw conservative external upper and lower bounds on *f* analogous to those given above for NYC. The lower bound is *f* = *D*_∞_/*Nr*_∞_ > *D*_∞_/*N* > *D*(July 9)/*N* = 0.09%. The upper bound is the ratio of the number of deaths to the number of reported cases; from the Illinois Public Health website, as of mid-July this ratio was 4, 776/98, 670 = 4.8%. This could be strengthened using an analysis similar to the case data lower bound for NYC, but the bound would still be well above the preferred model parameter region for *f*, as shown in the upper right panel of fig 12. This is shown more quantitatively in Fig. 11, which shows that the model with *f* = 1.1% predicts case numbers well above the reported cases.

**Figure 11:**
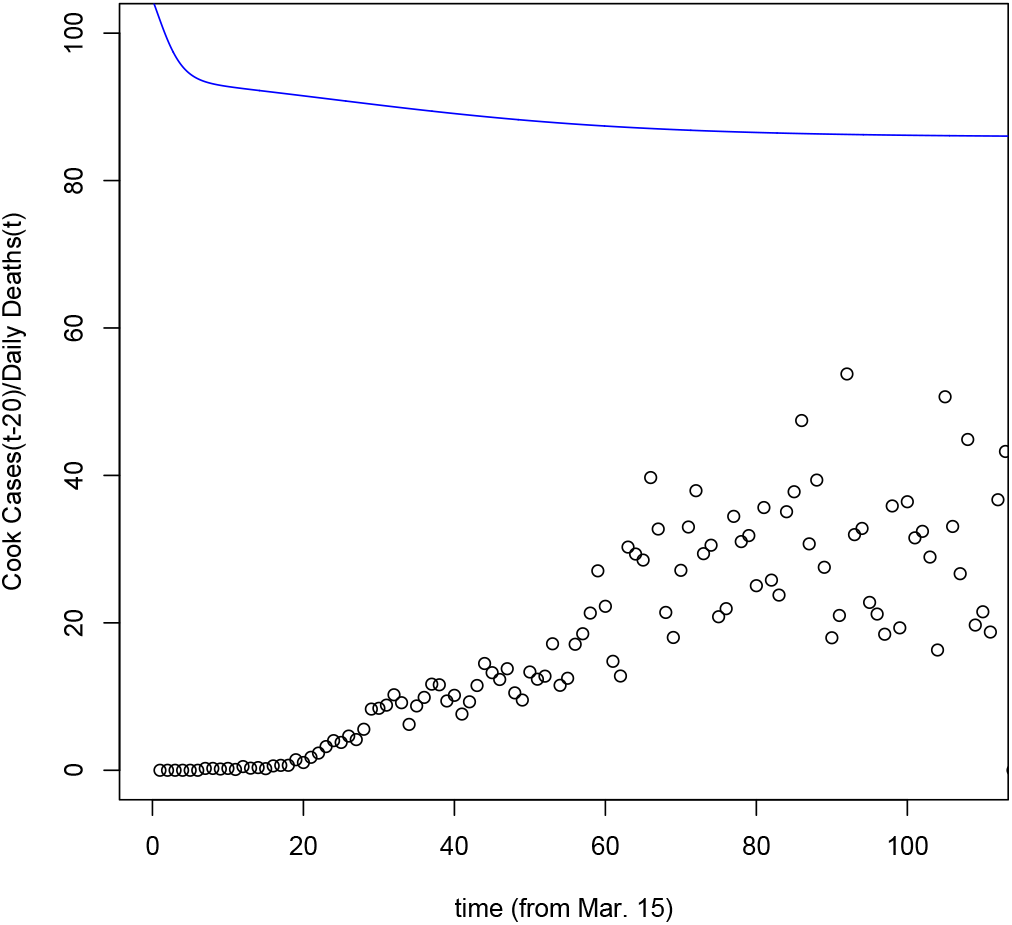
Ratio of time-shifted, Cook County case data to fatality data, (*dC*/*dt*)*_d_*(*t* − 20)/(*dD*/*dt*)*_d_*(*t*), the RHS of Eqn. (16) (points). Blue curve shows *R_t_s*/ *f* for a Cook County model with the lowest value of this quantity, with *R*_0_ = 1.24, *X* = 16.5%, and *f* = 1.1% (bottom row of Table 2 and Fig. 12). The model cases lie well above the reported cases; for the other models in Table 2, the curves would be even higher. While the case-to-fatality data ratio looks similar to that of NYC (compare left panel of Fig. 4), the models indicate that Cook County reported-to-actual case ratio is lower than that of NYC.

**Figure 12:**
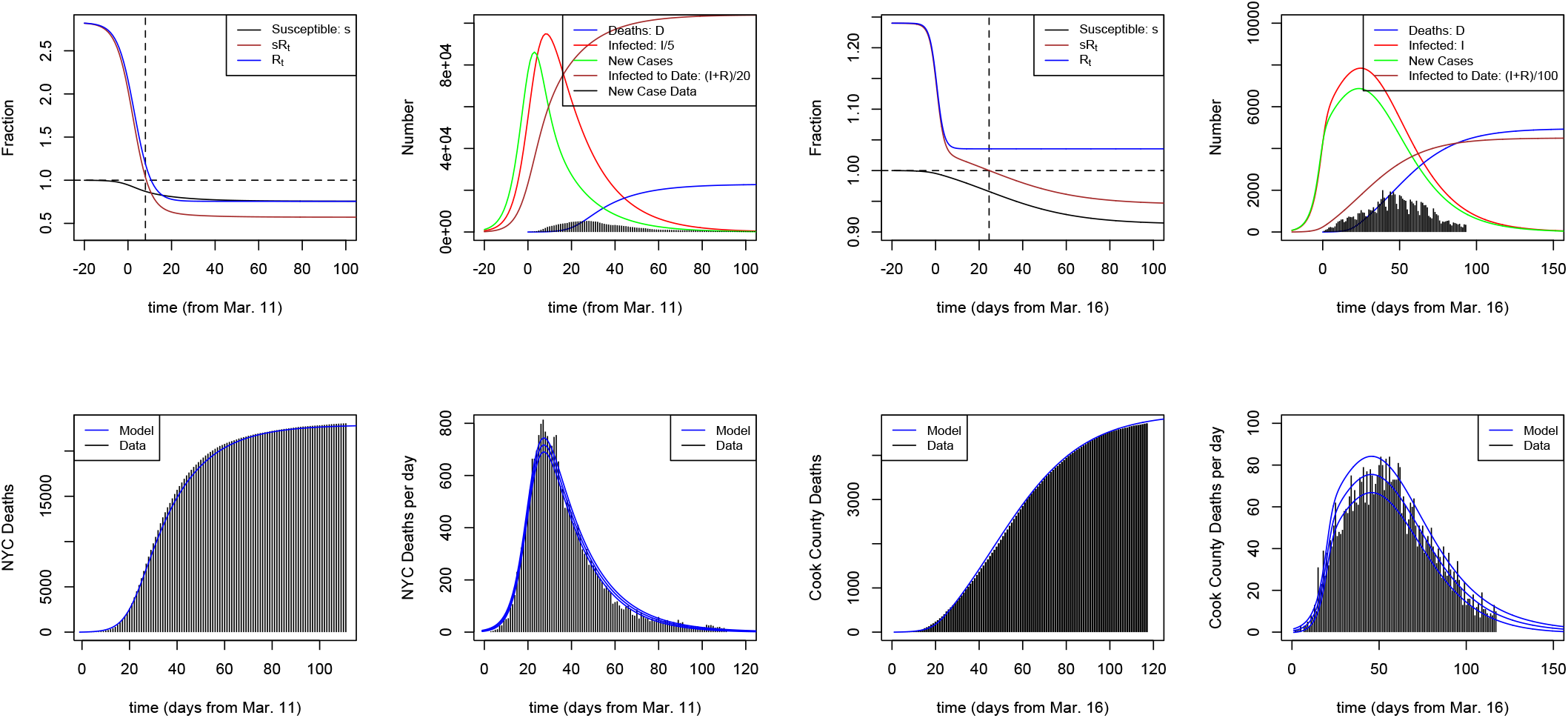
*Left panels:* Best-fit model for NYC with fixed *f* = 1.1% (2nd row of Table 2), with *R*_0_ = 2.82 and 100*X* = 73.4% reduction in social contact; 24.3% of the population get infected. *Right panels:* Best-fit model for Cook County with *f* = 1.1% (bottom row of Table 2), with *R*_0_ = 1.24, 100X = 16.5%, and 8.7% asymptotically infected.

## 5 Discussion: NYC vs. Cook County

The SIR model with a single transition in social contact in mid-March appears to provide a qualitative description of the fatality evolution in NYC (in terms of goodness of fit) and a quantitative description for Cook County, and can be used to get a qualitative understanding of the different behaviors of the data in the two places. NYC fatalities rose and fell sharply, with a FWHM of daily fatalities of about 20 days. This is attributable to a steep decline in the reproduction parameter *Rt*: the reduction in social contact was 74% in the best-fit models. By contrast, for Cook County *R_t_* appears to have fallen more modestly to values just above unity, leading to an extended plateau in daily fatalities, with a FWHM of order 55 days.

Despite the degeneracies among the model parameters, the qualitative differences in fatality evolution in the two urban areas point to their occupying different regions of model parameter space. As Table 2 shows, while the inferred values of *R*_0_ are comparable for the two locations, the best-fit values of the contact reduction factor, *X*, and the infection fatality rate, *f*, appear to be significantly higher for NYC than those for Cook County, while the asymptotic reproduction number, *R*_∞_, and the asymptotic fraction of population infected, *r*_∞_, appear lower. Since similar mitigation measures were instituted in both regions, it is not immediately obvious why NYC would have achieved a greater percentage reduction in social contact.

As for the significantly higher IFR value for NYC, there are numerous factors that might have contributed to this difference, if it is real, including: (1) differences in demographics and fractions of at-risk population, (2) availability of treatment, e.g., number of intensive care unit beds relative to population size, (3) hot spots in at-risk subpopulations, e.g., long-term care facilities, prisons, etc., (4) possible differences or evolution in lethality of COV-SARS-2 strains circulating in the two regions, (5) differences between documented population, *N*, and effective model population, *N_eff_*. On the last point, a critical simplifying assumption of the model is that every individual in the documented population *N* of each urban area is a ‘participant’ in the model, in the sense that they have equal non-zero probability of infection or transmission. However, if a subset *N* − *N_eff_* of the population has no social contact over an extended period, in principle they should be removed from the model population, reducing *N* to *N_eff_*. Since the IFR only enters the model through the combination *fN*, the true value of *f* would be higher than the estimate given in Table 2 by the factor *N*/*N_eff_*. For example, for *N_eff_* = *N*/2, the best-fit model estimate for *f* would double. Since roughly half of the Cook County population lives in suburban areas, with lower population density than the city of Chicago, one might expect it to have a larger value of *N*/*N_eff_* than for NYC; on the other hand, case and fatality numbers for the city and the suburbs appear comparable. In any case, this difference does not by itself appear to account for the nearly order of magnitude difference in the estimated values of *f* between the two regions.

Interestingly, the model parameter degeneracies in the two regions show both similarities and differences. In both regions, *R*_0_ and *X* are strongly positively correlated, and in both regions the fact that *D*_∞_ is well determined implies a tight inverse relation between *r*_∞_ and *f*, as shown in Table 2. However, for NYC, *X* (or *R*_0_) and *f* are positively correlated (see Fig. 6), while for Cook County they are strongly and tightly negatively correlated (see Fig. 9); also, for NYC, *R*_0_ and *R*_∞_ are negatively correlated, while for Cook County they are positively correlated. For Cook, unlike NYC, the contact reduction parameter *X* appears to ‘saturate’ at a value of about 55%, well away from the boundaries of the prior.

This difference in parameter degeneracies between *R*_0_ and *f*, as well as the differing constraints from case data, have important implications for the model parameter credible ranges for the two places. For models that fit the data, we have *i*(*t*)=(*dD*(*t* + Δ*t*)/*dt*)(*R*_0_ − 1)/*fNc*, where *c* = 0.25(0.21) for NYC (Cook). The coefficient of the *dD*/*dt* term in this expression varies by only ~ 30% over the 95.45% credible interval of model parameters for NYC, but it varies by nearly an order of magnitude for the corresponding parameter range for Cook County. In other words, the NYC fatality data constrain the model curve of infections, *i*(*t*), to lie within a narrow range in amplitude, e.g., the peak in number of individuals infected is not far from 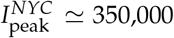, while for Cook County a much broader range in the amplitude of *i*(*t*) can reproduce the *dD*/*dt* data, with 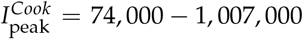 at 95% CL. This is reflected in the larger range of allowed parameter values for Cook, particularly for *r*_∞_.

From Table 2, the asymptotic model projection for NYC is about 270 deaths per 100,000 population, while for Cook County it is only about 100. Yet as just noted, the inferred value of *R*_∞_ for NYC appears lower than for Cook County and its contact reduction factor, *X*, is correspondingly higher. Other things being equal, we would expect larger social contact reduction to lead to fewer fatalities. In the model, this counterintuitive difference is due to the much lower IFR for Cook. Recall that the asymptotic fatalities per population are given by *D*_∞_/*N* = *fr*_∞_. While the estimated *r*_∞_ for Cook is higher than for NYC, this is more than compensated by the lower estimated value of *f*, resulting in the 2.7 ratio for *D*_∞_/*N* values between the two places. According to the model, early in the pandemic *D*(*t*)/*N* for NYC rose more rapidly than for Cook due to (a) the larger exponent *γ*(*R*_0_ − 1) for growth and (b) the inference that mitigation measures in Cook County were imposed ~ 2 days earlier and more rapidly, relative to the time of initial outbreak, than in NYC. Accordingly, the ratio of *D*(*t*)/*N* values peaked at a value of 8.3 before declining toward its asymptotic value of 2.7. This difference in ‘response times’ is seen in the upper panels of the preceding model Figures: for Cook County, *R_t_* reached its asymptotic value *R*_∞_ well before the peak in the number of individuals infected occurred, while for NYC *R_t_* was still falling when infections peaked. An important note of caution regarding differences in *D*/*N* is that Cook County reported deaths are likely a smaller fraction of true deaths than NYC Public Health reported deaths, since the Cook County Medical Examiner does not appear to include probable COVID-related deaths in its public database. For NYC, about 20% of the total reported deaths are classified as probable, and I have included them in the data without differentiation from confirmed deaths.

As Table 2 indicates, the credible ranges of values for *f* for NYC and Cook County are surprisingly different from each other, particularly given the similarity in the ratio of reported case to fatality data; in particular, the inferred values for *f* and for *r*_∞_ seem respectively low and high for Cook County. As Table 2 also indicates, the fits in both locations with fixed values of *f* away from the best-fit models are not dramatically worse—the likelihood is a weak function of *f*, particularly for Cook County—which suggests that even a modestly informative prior on *f* would shift the results. Suppose, for example, that external information indicated *f* = 1.1% in both locations. This would flip the above interpretation of the NYC-Cook difference in *D*_∞_/*N*: it would be attributed not to the difference in *f* but to the fact that a 2.7 times *higher* (not lower) fraction of those in NYC were infected than in Cook, 24 vs. 9%. In this case, one would need to explain not the large difference in *f* but the large difference in *X*, 73 vs. 17%, and in mean removal time *γ*^−1^, 7.3 vs. 1.1 days. Fig. 12 shows the best-fit models for both locations with *f* = 1.1%.

Finally, as noted above, the goodness of fit measures *RD_MLE_* of the MLE NYC and Cook County models differ significantly from each other. Fig. 13 shows the cumulative contributions to *RD_MLE_* for each location. For Cook County, the trend is roughly linear in time, as would be expected. For NYC, roughly half of the contribution to *RD_MLE_* comes from the relatively short periods before 15 days and after 100 days, suggesting that those periods are less well described by the model; for the intermediate period, *RD ≃* 200 for about 85 degrees of freedom for NYC.

**Figure 13:**
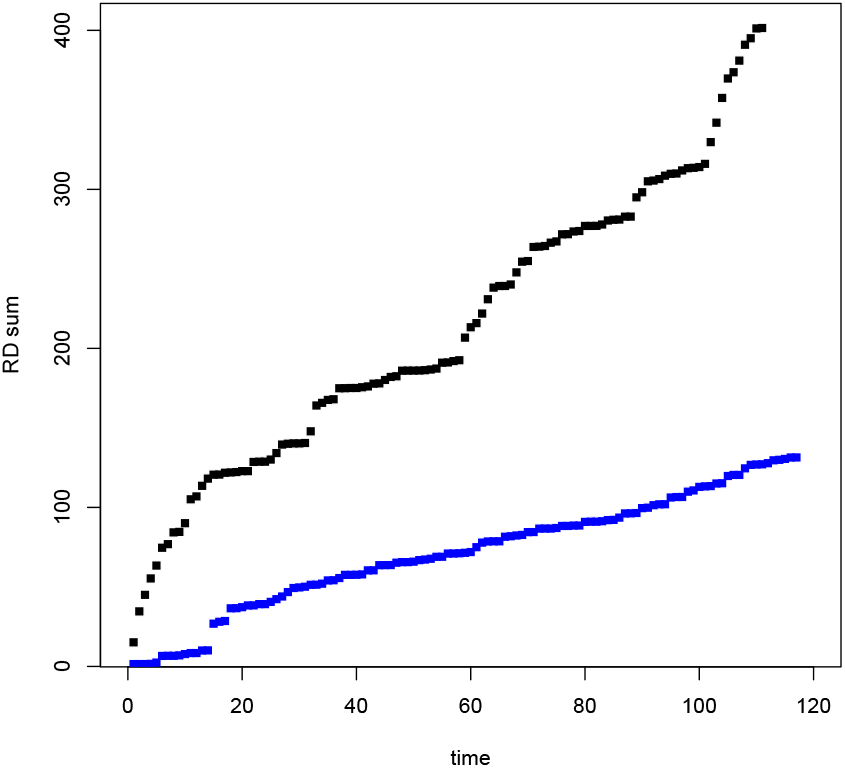
Cumulative contributions vs. time to the goodness of fit measure, *RDMLE*, for the MLE models for NYC (black points) and for Cook County (blue points).

## 6 ‘Re-opening’: Easing of Contact Reduction Measures

In the previous sections, I assumed that mitigation measures taken in mid-March in NYC and Cook County remained in place through early/mid June, which is consistent with the fatality data to the end of June in both locations. If they continue to remain in place in until a vaccine or effective treatment becomes available, then the pandemic should remain under control in those two areas. However, at this writing (mid-July), other parts of the country have ‘re-opened’, leading to a surge in U.S. cases that could potentially seed new outbreaks in NYC and Cook County. In addition, both urban areas have begun and are planning further steps in phased processes of easing of mitigation measures that are likely to result in increasing the reproduction number, *Rt*, above its current value. In this section, I consider the potential impacts of such easing on the future course of the pandemic in NYC and Cook County, adopting the same model assumptions as above, including continued constancy of *f* and *γ* over time.

I model this ‘re-opening’ by assuming that at a time *t_e_* > *t_m_*, social contact is increased by a factor *X_e_* over a time period *ω*^−1^, which introduces 3 additional parameters into the model for *R_t_*, i.e.,

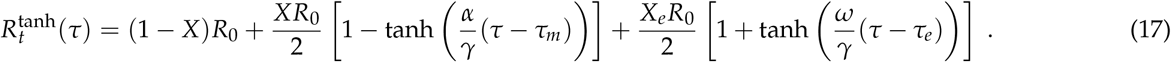

In this model, *R_t_* decreases from its initial value, *R*_0_, to (1 − *X*)*R*_0_ around time *t_m_* and then increases to 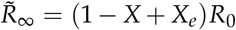 around time *t_e_*, where in this section 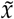 denotes the asymptotic value of parameter *x* after the 2nd wave. Assuming *t_e_* ≫ *t_m_*, at times *t_m_* ≪ *t* ≪ *t_e_* the dynamical *SIR* variables will have approximately reached their asymptotic values from the initial mitigation stage, e.g., *r ≃ r*_∞_, where *r*_∞_ values are given in Table 2, before subsequently evolving due to the easing of measures. An approximate condition for the disease to again become pandemic is therefore 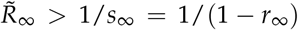, which implies a second wave will occur if

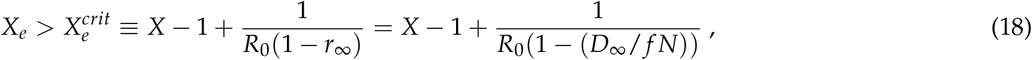

where again *D*_∞_ is the asymptotic model fatality count at the end of the first wave. The corresponding values of 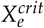 are given in Table 2.

For the NYC best-fit models (first row of Table 2), the value of 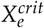 is tightly constrained to about 19%, nearly independent of the parameter degeneracies; this is because the condition 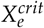 traces out a locus in the *R*_0_-*X* parameter space, *X* − 1 = 0.19 − 1/(*R*_0_(1 − *r*_∞_)), that is close to that in the right panel of Fig. 4. For NYC, this value of 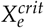 is relatively small compared to the values of the initial contact reduction factor *X* ≿ 0.7 for the corresponding models, which means that only modest re-opening measures are likely to trigger a second wave. However, the severity of the 2nd wave will depend on *f* and on *X_e_*.

**Figure 14:**
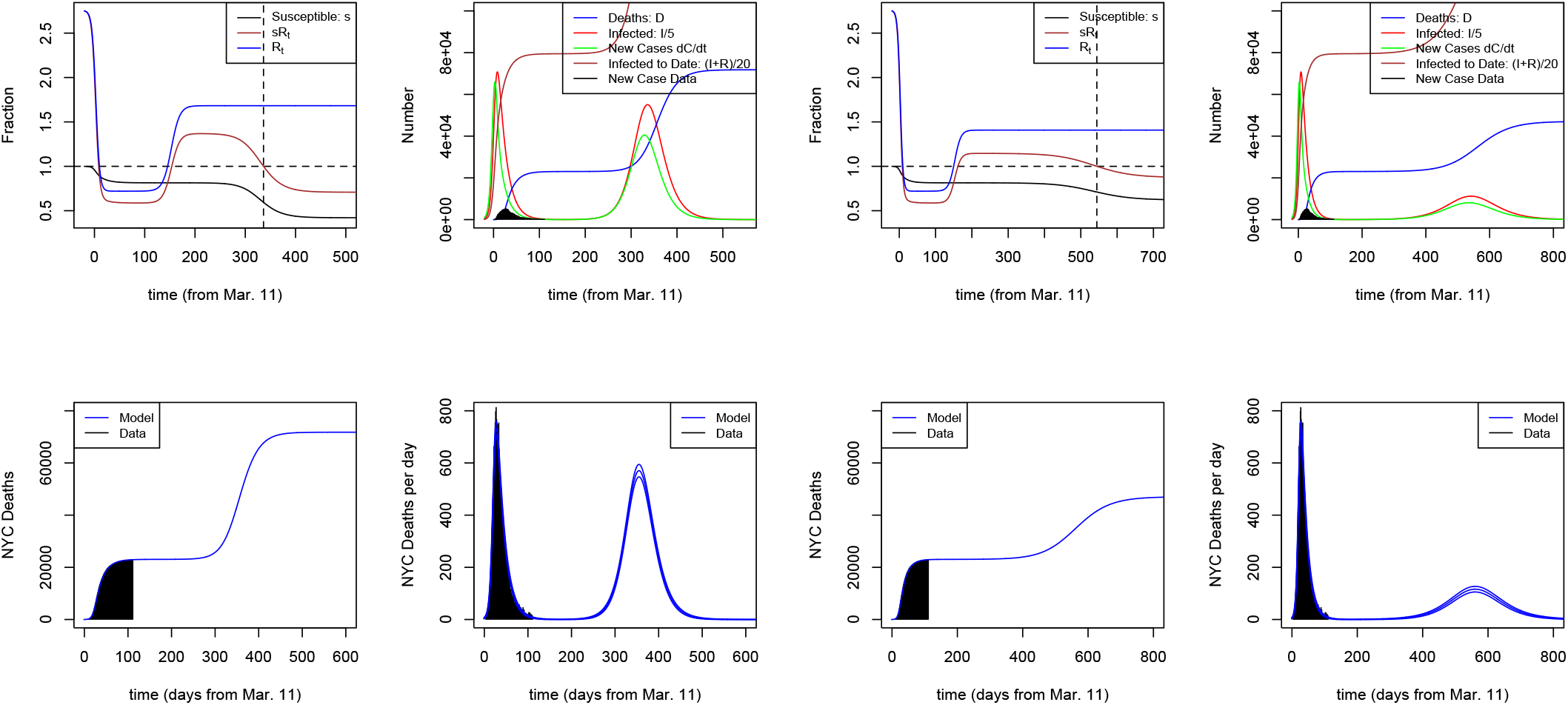
*Left panels:* SIR model of Fig. 5 (2nd row of Table 2) for NYC, now with 100*X_e_* = 35% increase in social contact phasing in over 17 days in mid to late August 2020, leading to a large 2nd wave that peaks in early 2021. *Right panels:* Best-fit model for *f* = 1.1% (3rd NYC row of Table 2 and Fig. 12) followed by re-opening (contact increase) with 100*X_e_* = 25%

Fig. 14 illustrates this. The left panel starts with the best-fit model of Fig. 5 (1st row of Table 2) and introduces an easing of mitigation parameterized by *X_e_* = 0.35 over an 1/*ω* = 17-day transition period with mid-point at *t_e_* = *t_m_* + 150 days, or Aug. 11, which yields 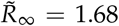. Daily fatalities bottom out in late August 2020 before ticking up. In this example, new cases peak in Feb. 2021, and daily fatalities peak around 550 per day in Mar. 2021. Here, with *f* = 1.45%, asymptotically 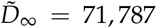, and 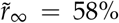 of the NYC population ends up infected. (This model makes the unrealistic assumption that mitigation measures would not be reinstituted once the second wave starts or alternatively demonstrates a possible outcome if they are not reinstituted.) By contrast, the right panel of Fig. 14 implements easing of mitigation with *X_e_* = 0.25 (same values of *t_e_*, *ω* as above), starting from the best-fit NYC model with fixed *f* = 1.1% (2nd row of Table 2), which yields 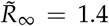, 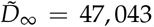, and 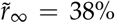 asymptotically infected. In the left (right) panel 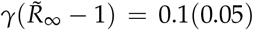 inverse days, so the 2nd-wave growth rate is twice as fast in the left panel. With its larger *X_e_*, the left-hand case yields 1.5 times as many fatalities as the right-hand, even though the IFR is only 1.3 times as large. To keep future fatalities under control as NYC reopens it will be imperative either to keep the easing below 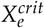 or to take other measures to drive down the IFR through improved treatment and/or extra protections for at-risk subpopulations, as well as widespread and *rapid* testing and quarantine to reduce the mean removal time *γ*^−1^.

For Cook County, there is much wider variation in the values of 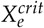 within the family of best-fit models (those in the first row of Cook County entries in Table 2). As a result, it is much more challenging to set acceptable reopening thresholds for Cook County. However, for values of *f* at the upper end of the allowed range for Cook County, the re-opening threshold for triggering a 2nd wave is quite low, of order 10% increase in social contact (last two rows of Table 2). As an example, Fig. 15 shows a re-opening scenario for Cook County, starting from the *f* = 0.4% model of Fig. 8 (2nd row of Cook entries in Table 2). Here, *X_e_* = 0.18, so the increase in social contact is half of the initial *X* = 0.36 reduction, with the re-opening taking place over 17 days centered on *t_e_* of mid-July. Since 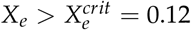 for these model parameters, there is a second wave in which fatalities peak in December; while flatter than the first wave, it nevertheless yields *D*_∞_ = 7291, a significant increase compared to the present. On the other hand, for the MLE model of Fig. 7, the same re-opening parameters would not cause a second wave, since 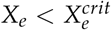.

**Figure 15:**
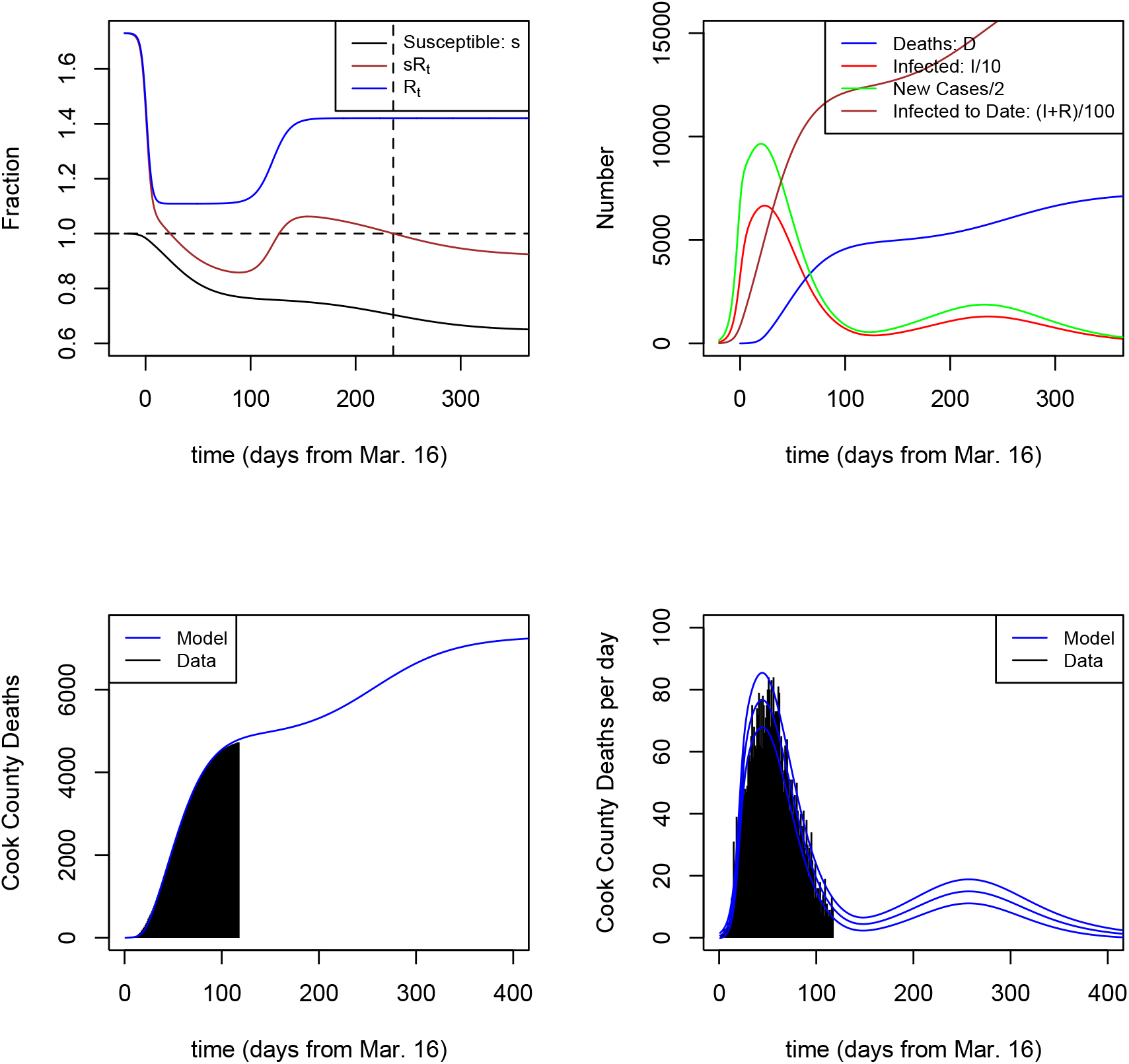
SIR model of Fig. 8 (2nd row of Cook entries in Table 2) for Cook County, with 100*X_e_* = 18% increase in social contact phasing in over 17 days centered in mid-July, leading to an extended 2nd wave. For the MLE model (first row of Cook entries in Table 2), the same reopening does not trigger a 2nd wave, since 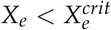.

The important conclusions from this study are that the impact of reopening measures depend critically on (a) whether they are ‘above threshold’, i.e., 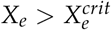, (b) the value of the IFR, *f*, and (c) the removal timescale, *γ*^−1^, which impacts the 2nd-wave growth rate.

## 7 Conclusion

I draw several conclusions from this modeling exercise: (1) *SIR* models with a single transition to reduced social contact in mid-March appear to describe the fatality data to the end of June for NYC at least qualitatively and for Chicago quantitatively; (2) despite significant parameter degeneracies, the model for NYC became quasi-predictive in the sense that model fits made soon after fatalities started to decline accurately predicted the subsequent month’s evolution of fatalities; (3) the evolution of the “first-wave” pandemic differed markedly between the two locations, pointing to different values of the model parameters—the public health reasons behind these differences would be interesting to explore; (4) only modest easing of contact reduction measures (‘re-opening’) may be needed to trigger a second wave in either city, in the absence of other measures; (5) the threshold for ‘safe’ reopening appears well-constrained by the model for NYC but not for Cook; and (6) the impact of such easing will depend critically on the amplitude of contact increase, *X_e_*, the infection fatality rate, *f*, and the mean removal time from the population, *γ*^−1^.

This document was prepared using computing resources of the Fermi National Accelerator Laboratory (Fermilab), a U.S. Department of Energy, Office of Science, HEP User Facility. Fermilab is managed by Fermi Research Alliance, LLC (FRA), acting under Contract No. DE-AC02-07CH11359.

## Data Availability

Based upon publicly available fatality data at the sites below.

https://nychealth/coronavirus

https://maps.cookcountyil.gov/medexamcovid19/

## A Critical Slowing Down in the *SIR* Model

For the SIR model, the condition *R_t_s* = 1 marks a critical transition from growth to decay of the pandemic. The approach to critical transitions often involves a qualitative change in behavior of the system. The dynamics for Cook County shown in Figs. 7 and 8 shows a transition from exponential to roughly linear time dependence for *D*(*τ*). To understand when this is expected to occur, we can define new variables,

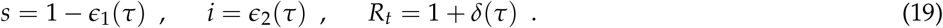

Here, *ϵ*_1_ and *ϵ*_2_ are the dynamical variables of the evolution, and *δ*(*τ*) is fixed by the mitigation model, e.g., Eqn. (8). Substituting these expressions into Eqns. (6), we find

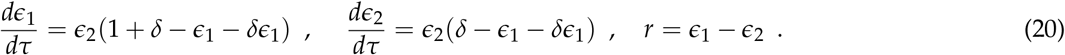

If the mitigation model *R_t_* is such that, over some time interval, *δ* ≃ *ϵ*_1_/(1 − *ϵ*_1_), then over that interval, *dϵ*_1_/*dτ* ≃ *ϵ*_2_, *dϵ*_2_/*dτ* ≃ 0, and *dr*/*dτ* = *ϵ*_2_ ≃ constant. Over this time interval, *s* decays linearly, *r* grows linearly, and *i* is approximately constant: the number of infected hits a plateau, which will lead to a plateau in daily deaths a time Δ*τ* later. This appears to be an example of the phenomenon of “critical slowing down” in the approach to equilibrium in second-order phase transitions as well as in certain dynamical systems: in this case, the time evolution transitions from early exponential behavior to linear behavior [12].

Note that I have not assumed that *ϵ*_1_, *ϵ*_2_ or *δ* are very small compared to 1. For the best-fit Cook County models (first Cook County row in Table 2), *δ* ≃ 0.07 − 1.2. For the mitigation model of Eqn.(8), *δ* is nearly constant over the time interval of interest, while *ϵ*_1_ is a growing function of time (*s* is monotonically decreasing), so this quasi-equality will only last for a limited time interval, given our assumptions. Since *ϵ*_1_ grows linearly during this stage, 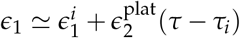, the characteristic length of the plateau stage in real time should be of order 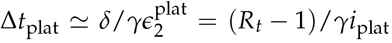, where *i*_plat_ is the value of *i* at peak. This is about 30 − 35 days for the models in Fig. 7 and 8.

## B Semi-Empirical Model

In the body of the paper, I adopt and test the assumption that social contact reduction measures in both locations were implemented in a single monotonic transition of limited duration, after which social contact rates were stable in the population. The analysis led to inferences about the parameters describing this transition. An alternative, discussed here, is to adopt a semi-empirical approach that infers features of the model directly from the fatality data to the extent possible. The advantage of the semi-empirical approach is that it does not make the above assumptions about monotonic behavior between two asymptotic regimes of constant social contact. The disadvantages of the approach are that (a) it is not informed by public information about mitigation measures, which led to the hypothesis of Eqn. (8), (b) since it infers the *SIR* functions directly from the fatality data (up to unknown model parameters), it does not allow one to project the model forward in time, i.e., to make predictions, and (c) in the absence of other data or priors, it does not enable one to derive constraints on the parameters *f*, *γ*, and *R*_0_: the data map onto models with arbitrary values of these parameters.

From Eqns. (5) and (6), we can write the *SIR* model functions in terms of the fatality data as

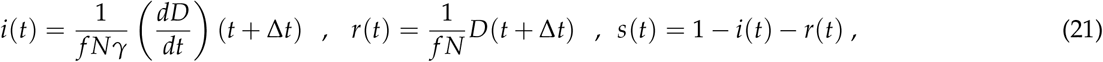

and we can infer the time-dependent reproduction number via, e.g.,

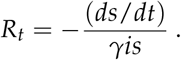

Since the latter involves differentiation of the noisy daily fatality data, a suitable smoothing over a several-day window can be applied. The semi-empirical approach is thus a model with *k* + 2 parameters, where *k* is the number of days of fatality data, and *f* and *γ* are the two additional parameters. This model therefore achieves no data reduction in the usual sense of the term.

As an example of the semi-empirical approach, Fig. 16 shows the model of Eqns. (19) and (20), with five-day smoothing of *R_t_*, for *f* = 4.9% and *γ*^−1^ = 16.6 days, the same values as for the NYC tanh MLE model (first row of Table 2), superimposed on the tanh MLE model (dashed lines).

**Figure 16:**
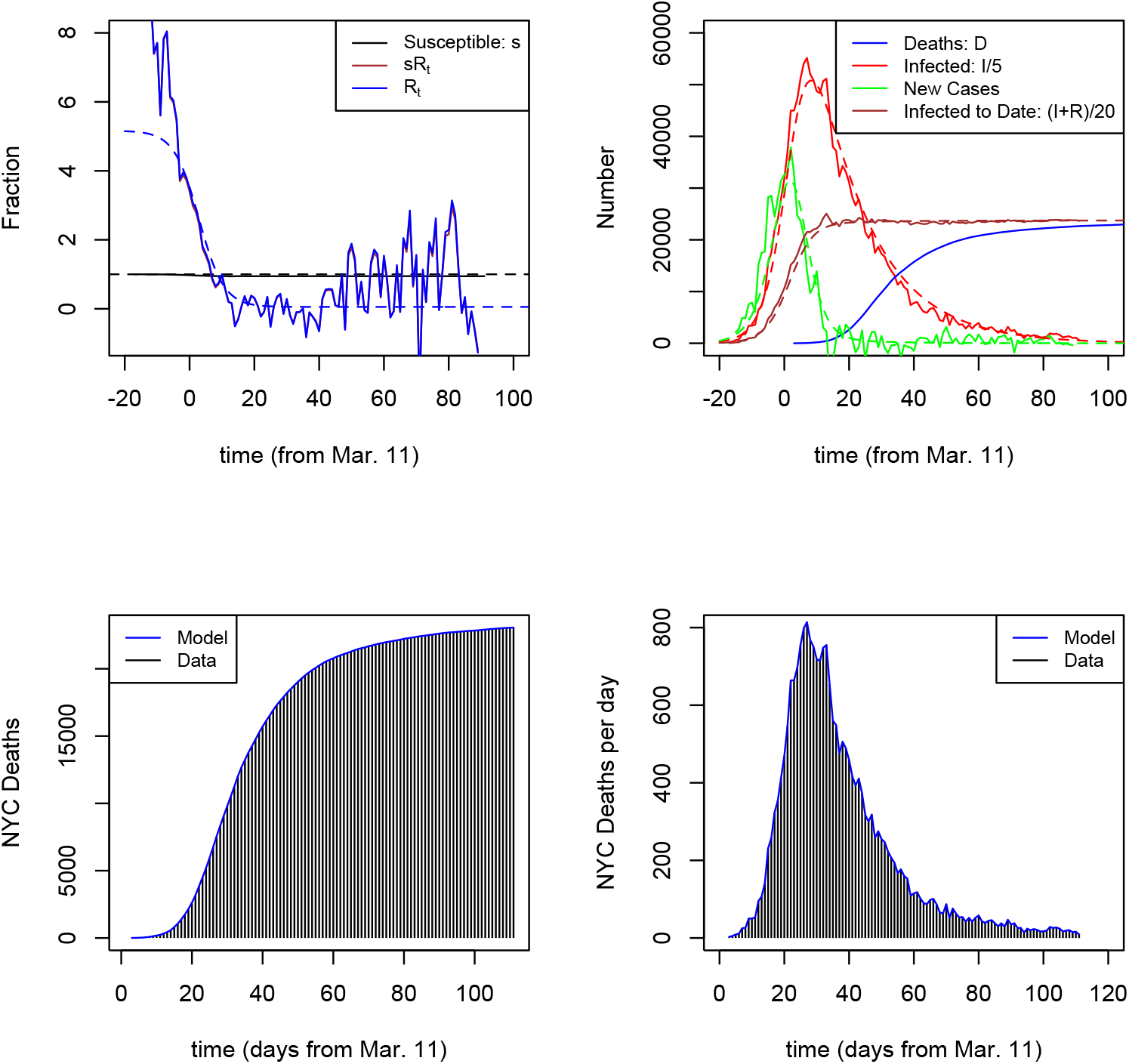
Semi-empirical model for NYC with same values of *f* and *γ* as the tanh MLE model of Fig. 3, *f* = 4.9% and *γ*^−1^ = 16.6 days, superimposed on the tanh MLE model (shown as dashed lines). For plotting purposes, the inferred *R_t_* has been filtered through a five-day window. Lower panels illustrate that this approach uses the daily and cumulative fatality data as input, as opposed to forward modeling of the data.

## References

1. P. E. Lekone and B. F. Finkenstadt, “Statistical Inference in a Stochastic Epidemic SEIR Model with Control Intervention: Ebola as a Case Study,” Biometrics, 62, 1170–1177 (2006).

2. S.-m. Jung, et al., “Real-Time Estimation of the Risk of Death from Novel Coronavirus (COVID-19) Infection: Inference Using Exported Cases,” J. Clin. Med., 9, 523 (2020).

3. N. Linton, et al., “Incubation Period and Other Epidemiological Characteristics of 2019 Novel Coronavirus Infections with Right Truncation: A Statistical Analysis of Publicly Available Case Data,” J. Clin. Med. 9, 538 (2020).

4. C. Branas, et al., “Flattening the Curve before it Flattens us: Hospital Critical Care Capacity Limits and Mortality from Novel Coronavirus (SARS-Cov2) Case in U.S. Counties,” medRxiv preprint 2020.04.01.20049759

5. W. Kermack and A. McKendrick, Proc. Roy. Soc. London A, 115, 700 (1927).

6. E. Beck and B. Armbruster, “Elementary Proof of Convergence to the Mean-Field Model for the SIR Process,” arXiv, 1511.08572.

7. J. Katz, D. Lu, M. Sanger-Katz, New York Times, April 28, 2020.

8. J. Wu, A. McCann, J. Katz, E. Peltier, New York Times, April 30, 2020.

9. F. Baty, et al., J. Stat. Software, 66, 1 (2015),

10. K. Soetaert, T. Petzoldt, and R. W. Setzer, J. Stat. Software, 33, 1 (2010).

11. W. H. Press, S. A. Teukolsky, W. T. Vetterling, and B. P. Flannery, “Numerical Recipes, the Art of Scientific Computing,” 3rd edition, Cambridge University Press, 2007.

12. M. Scheffer, et al., “Early Warning Signals for Critical Transitions,” Nature Reviews, 461, 53 (2009).

